# Epigenetic Biomarkers of Socioeconomic Status are Associated with Age-Related Chronic Diseases and Mortality in Older Adults

**DOI:** 10.1101/2024.05.21.24307701

**Authors:** Lauren L. Schmitz, Lauren A. Opsasnick, Scott M. Ratliff, Jessica D. Faul, Wei Zhao, Timothy M. Hughes, Jingzhong Ding, Yongmei Liu, Jennifer A. Smith

## Abstract

Later-life health is patterned by socioeconomic influences across the lifecourse. However, the pathways underlying the biological embedding of socioeconomic status (SES) and its consequences on downstream morbidity and mortality are not fully understood. Epigenetic markers like DNA methylation (DNAm) may be promising surrogates of underlying biological processes that can enhance our understanding of how SES shapes population health. Studies have shown that SES is associated with epigenetic aging measures, but few have examined relationships between early and later-life SES and DNAm sites across the epigenome. In this study, we trained and tested DNAm-based surrogates, or “biomarkers,” of childhood and adult SES in two large, multi-racial/ethnic samples of older adults—the Health and Retirement Study (HRS) (N=3,527) and the Multi-Ethnic Study of Atherosclerosis (MESA) (N=1,182). Both biomarkers were associated with downstream morbidity and mortality, and these associations persisted after controlling for measured SES, and in some cases, epigenetic aging clocks. Both childhood and adult SES biomarker CpG sites were enriched for genomic features that regulate gene expression (e.g., DNAse hypersensitivity sites and enhancers) and were implicated in prior epigenome-wide studies of inflammation, aging, and chronic disease. Distinct patterns also emerged between childhood CpGs and immune system dysregulation and adult CpGs and metabolic functioning, health behaviors, and cancer. Results suggest DNAm-based surrogate biomarkers of SES may be useful proxies for unmeasured social exposures that can augment our understanding of the biological mechanisms between social disadvantage and downstream health.

**Significance Statement:** Information on DNA methylation (DNAm)—an epigenetic modification that plays a central role in regulating gene expression—is increasingly available in large epidemiological studies. Since DNAm is relatively stable but responsive to environmental influences, genome-wide signatures are promising surrogates or biomarkers of exposure that may both shed light on biological mechanisms between adverse environments and downstream health and/or act as proxies for unmeasured exposures. To better understand the biological embedding of social disadvantage, this study trained and tested DNAm-based surrogates of childhood and adult socioeconomic status (SES) in two US-based cohorts of older adults. Findings reveal distinct DNAm signatures of SES that connect social adversity across the lifecourse with dysregulated immune system responses, inflammatory pathways, poorer metabolic functioning, chronic diseases, and cancer.

## Introduction

Health outcomes in later life are patterned in part by socioeconomic influences across the lifecourse (1, 2). However, the mechanistic pathways underlying the biological embedding of socioeconomic disadvantage and its consequences on morbidity and mortality are not fully understood. One potential pathway may be through modification of epigenetic mechanisms, such as DNA methylation (DNAm), that regulate gene expression. Alternatively, DNAm could be a marker of underlying biological response to environmental or contextual exposures. Recent studies have shown that socioeconomic status (SES) is associated with epigenetic aging measures (3, 4), but few studies have examined relationships between indicators of early and later-life SES and DNAm sites across the epigenome (5).

DNAm is the covalent addition of a methyl group to the 5^th^ carbon on a cytosine DNA base. In humans, DNAm occurs at sites on the genome where a cytosine is followed by a guanine, separated by a phosphodiester bond (CpG site). DNAm is the most well-studied type of epigenetic modification, and it is increasingly being profiled in large epidemiological studies. DNAm patterns are tissue-specific, influenced by both genetics and environmental exposures, and are relatively stable compared to other cellular biomarkers such as gene expression (6, 7). However, patterning at some CpG sites is dynamic throughout the life course and can shift in response to environmental stimuli; thus, it has been proposed as a possible mechanism linking social exposures to adverse health outcomes. More recently, epigenome-wide differences in DNAm have been associated with current and/or past environmental exposures, suggesting global variation in DNAm could serve as a cumulative or time-specific biomarker of exposure, regardless of whether each specific CpG site is mechanistically involved in the disease process (8–10). Thus, epigenetic signatures may be useful proxies of exposure data in epidemiology studies, particularly when collection of these data are costly and/or subject to recall bias (11, 12).

DNAm-based surrogates or biomarkers are typically constructed by regressing the exposure or health outcome of interest on a set of CpG sites using a supervised machine learning method (i.e., penalized regression) in a training sample. Currently, the most well-cited DNAm biomarkers are epigenetic aging measures or epigenetic “clocks” that are highly predictive of chronological age (13–20), biological age and mortality (21–23), and the rate of aging (24, 25). To date, over 30 epigenetic aging measures have been developed for adults and have been associated with numerous health, socioeconomic, and lifestyle factors (26–33). These methods have been extended to calculate surrogate DNAm biomarkers for hundreds of health outcomes and disease-associated plasma proteins, including cardiovascular disease, diabetes, C-reactive protein, HDL cholesterol, and interleukin-6 (34–37). In the case of health behaviors like smoking and alcohol consumption, DNAm surrogates are typically more accurate than self-reports, thereby reducing misclassification and improving disease prediction and risk stratification (38). For past exposures where retrospective data collection is difficult or even impossible to collect, DNAm-based surrogates hold more promise as an exposure biomarker than other omics data because DNAm is more chemically stable than RNA or metabolites and does not degrade as easily with long-term storage (12, 39). For example, DNAm biomarkers have been successful in reflecting past exposures to prenatal smoking, prenatal alcohol, maternal diet, exposure to lead, pesticides, and other toxins (40–42). Additionally, for some exposures like smoking, DNAm surrogates have shown more promise in accurately estimating exposure timing and dosage than prior gold standard smoking biomarkers like cotinine (12, 43).

With respect to SES, a DNAm-based surrogate may also be useful as a ‘discovery biomarker’ that can indicate biological processes that arise from or are associated with social disadvantage and its consequences. Thus, as opposed to more clinical biomarkers where disease prediction or risk stratification is the central goal (44), DNAm-based surrogates of SES may also help elucidate “biological expressions of social inequality”, or molecular pathways that link adverse social or environmental experiences with downstream health outcomes (45). To date, a large body of literature suggests that low SES is strongly associated with morbidity and mortality, including health problems such as cardiovascular disease, diabetes, hypertension, respiratory infections, and cancer (46, 47). Evidence suggests that the biological mechanisms linking SES with disease in industrialized populations operate through stress-related inflammatory pathways that can dysregulate immune system responses or metabolic functioning (48–50). In addition, mechanisms between socioeconomic adversity and health may vary depending on whether the exposure occurs in childhood, adulthood, or cumulatively over the lifespan. For example, the relationship between childhood adversity and later-life health may arise due to latent independent effects that disrupt critical or sensitive periods of development in early life (i.e., ‘biological embedding’ or ‘programming’ of adult disease), and/or via the role of early environment on subsequent life trajectories that have cumulative or ‘weathering’ effects on health (49, 51–54).

Epigenome wide association studies (EWAS) that independently test associations between individual CpG sites and SES-related outcomes generally corroborate these findings (5). However, more evidence is needed to understand the influence of timing and/or duration of socioeconomic disadvantage on DNAm in childhood and adulthood, as well as the degree of overlap in DNAm patterns across SES domains at both the individual and neighborhood levels (5). In addition, unlike penalized regression models that use shrinkage methods to construct DNAm biomarkers (e.g., lasso or elastic net), association weights from EWAS do not take the intercorrelation of CpG sites into account, which in turn may reduce the predictive accuracy of a biomarker or “methylation risk score” (MRS) that uses EWAS weights to calculate a weighted sum of DNAm levels (55).

In this study, we trained DNAm-based surrogates or biomarkers of childhood and adult SES in the Health and Retirement Study (HRS)—a large, multi-racial/ethnic sample of older adults (N=3,527)—and then tested their performance in another diverse, US-based sample of older adults (the Multi-Ethnic Study of Atherosclerosis or MESA, N=1,182). To capture the multidimensional nature of SES, DNAm biomarkers were trained on composite indices of childhood SES (cSES) and adult SES (aSES) that measure SES at both the individual and the sociological levels. We then examined whether these SES biomarkers (cSES-BIO and aSES-BIO) were associated with downstream morbidity and mortality, before and after accounting for self-reported childhood and/or adult SES and epigenetic aging measures. Finally, we explored potential biological mechanisms by functionally characterizing the individual CpG sites that comprise the SES biomarkers. Results indicate that biomarkers of SES across the lifecourse reveal additional information not captured in self-reported SES or next-generation epigenetic aging measures, suggesting they may not only be useful proxies for unmeasured social exposures, but they may also augment current knowledge about the biological impact of socioeconomic disadvantage on downstream health and aging.

## Results

### Sample statistics

The HRS analytic sample (N=3,527) had a mean age of 69.8 (SD=9.7) in 2016 (at the time of DNA methylation measurement) and was 58.7% female (*SI Appendix*, Table S1). Approximately two thirds of the sample (67.9%) were non-Hispanic white, with 15.6% non-Hispanic Black, 13.6% Hispanic, and 2.9% other. Most of the sample had a high school degree (81.5%), but only 25.6% had a college degree. Mean parental education was 10.9 years (SD=3.9), and mean childhood financial strain was 0.9 (SD=1.2). A total of 3,120 participants were followed-up and provided health information in 2018, and the sample statistics were substantively similar to those in 2016. The MESA replication sample (N=1,182) had a nearly identical mean age to the HRS sample (69.6, SD=9.3), but was slightly more diverse (White=48.5%; Black=19.4%; Hispanic=32.2%) and 50.9% female.

### SES index biomarkers

The analytic pipeline for biomarker creation and downstream analysis is shown in Figure 1 and described in detail in **Statistical Methods**. Due to the large number of CpG sites on the EPIC chip, we first conducted EWAS of the aSES and cSES indices separately in the training data (HRS) to reduce the number of CpGs evaluated by the elastic net procedure for biomarker construction. DNAm in HRS was profiled using the Illumina Infinium Methylation EPIC BeadChip. A total of 69,000 and 88,681 CpG sites on the EPIC chip were nominally associated (p<0.05) with aSES and cSES in the EWAS, respectively. Manhattan plots for these EWAS can be found in *SI Appendix*, Figures S1-S2.

**Figure 1.**
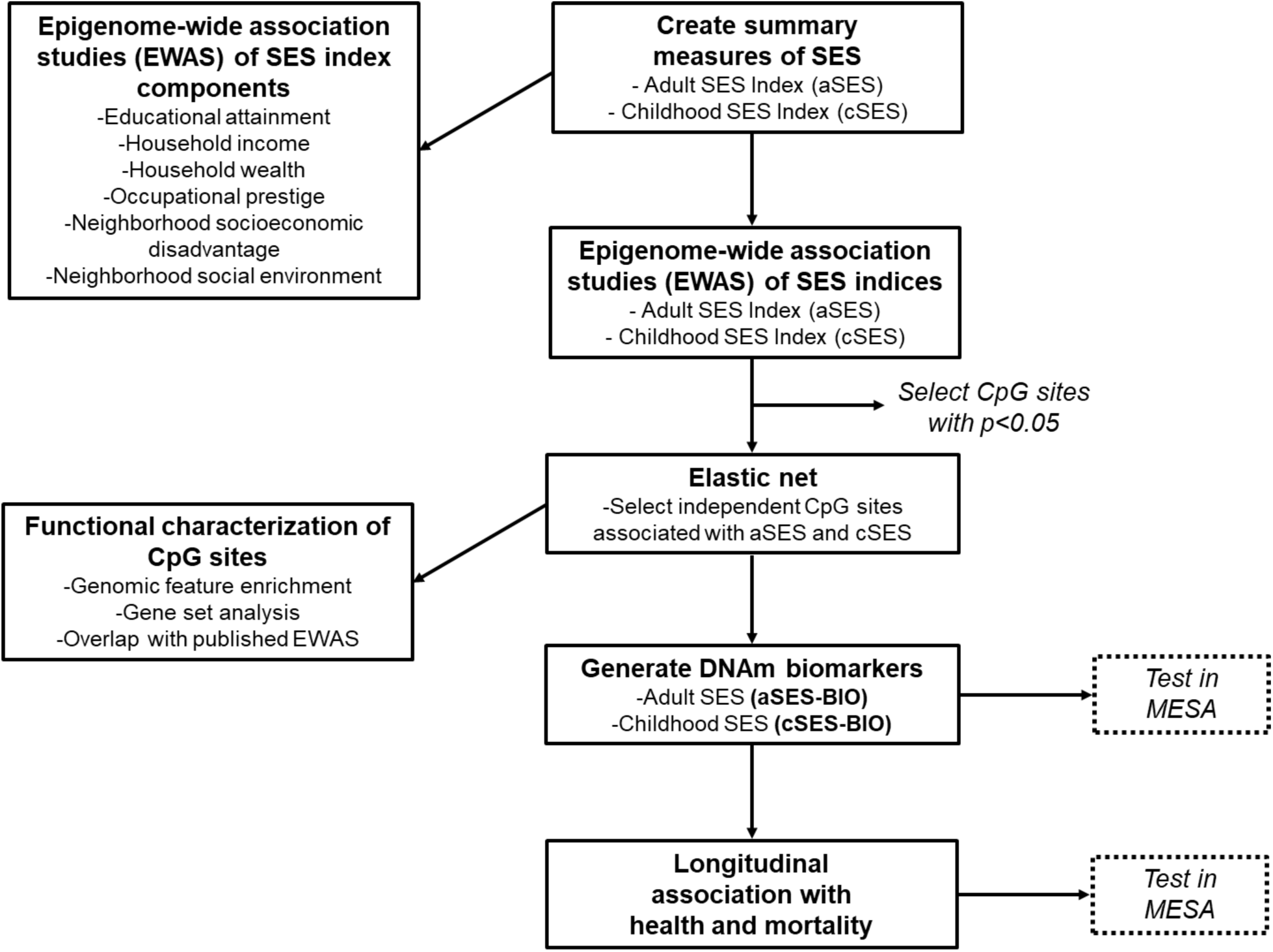
Analytic Pipeline in HRS and MESA

The elastic net procedure that minimized prediction error in the full sample selected a total of 25 CpG sites for aSES (lambda=0.138, cross validation (CV) R^2^=5.57%, Table 1) and 17 sites for cSES (lambda=0.128, CV R^2^=3.35%, Table 2). For both aSES and cSES, the selected CpG sites tended to have relatively high ranks in the original EWAS (ranks range from 1-15,540 for aSES and 1-43 for cSES, all with EWAS p-values < 4.6x10^-3^). EWAS of the individual components of the SES indices showed that education, household income, wealth, and neighborhood socioeconomic disadvantage had the strongest associations with many of the CpG sites that comprise aSES-BIO (*SI Appendix*, Table S2). Parental education had the strongest associations with many of the CpG sites that comprise cSES-BIO (*SI Appendix*, Table S3).

**Table 1.**
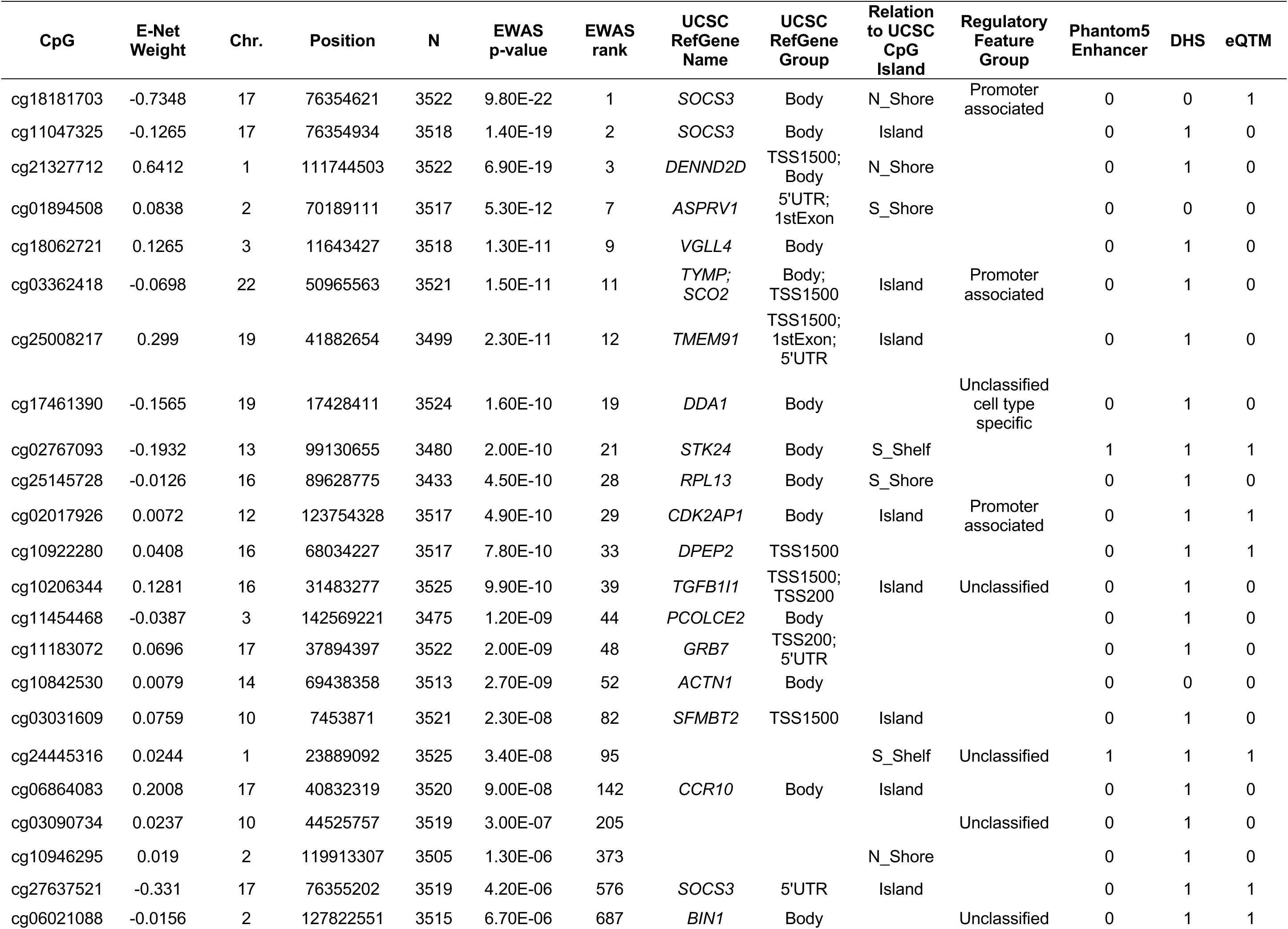
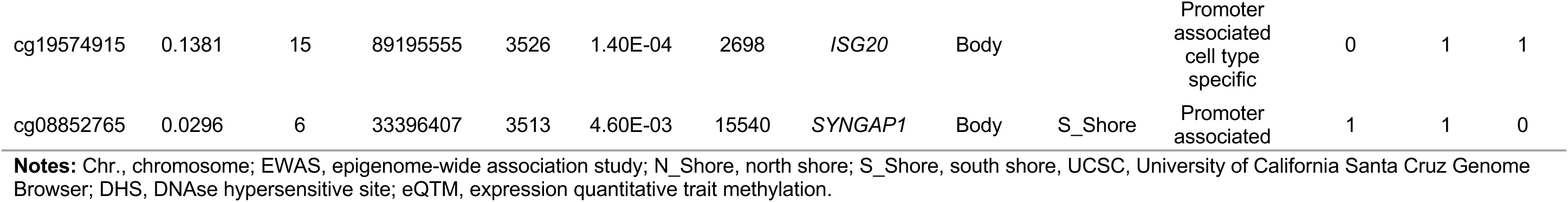
CpG sites selected for adult SES index biomarker (aSES-BIO) by elastic net in HRS.

**Table 2.**
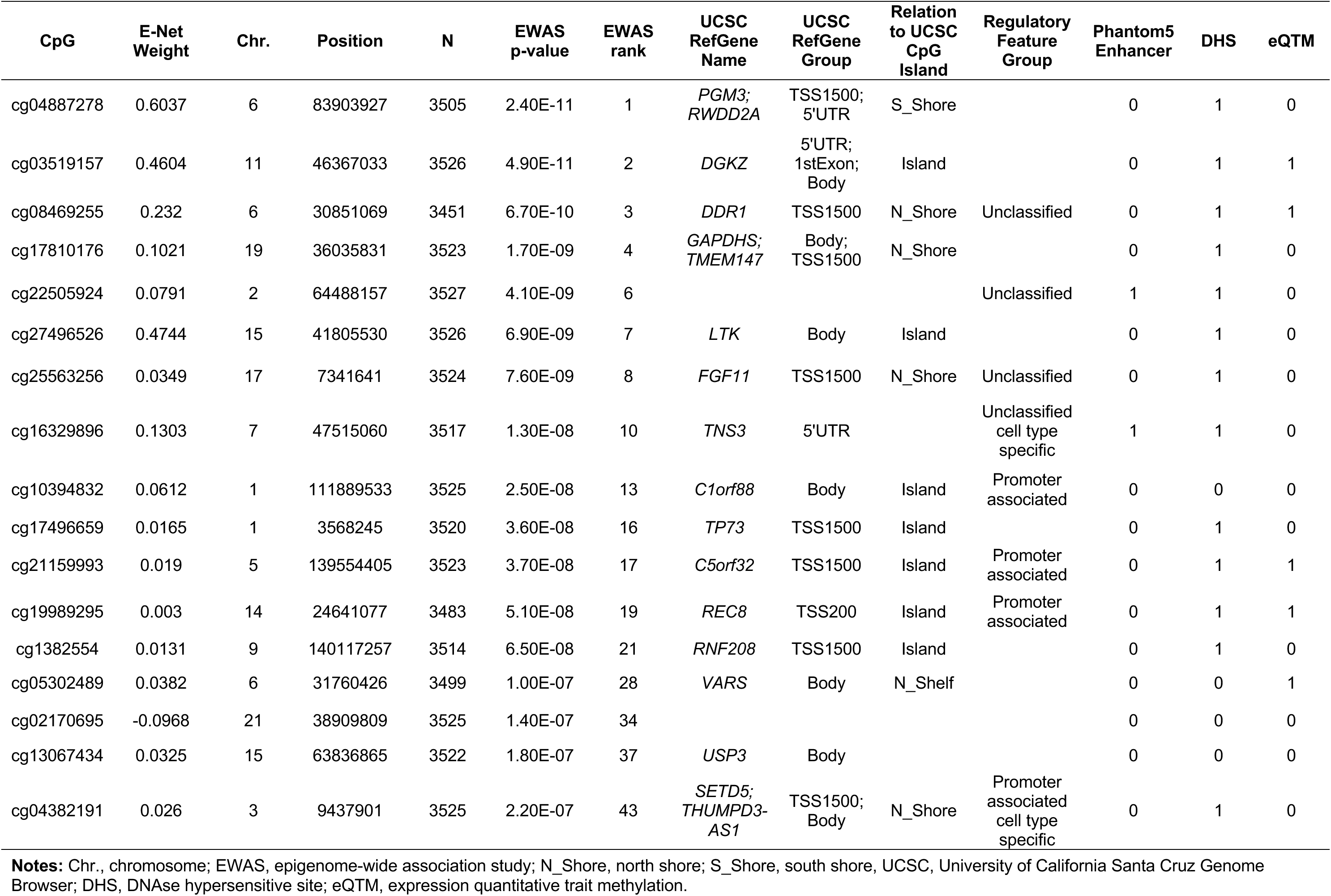
CpG sites selected for childhood SES index biomarker (cSES-BIO) by elastic net in HRS.

| CpG | E-Net Weight | Chr. | Position | N | EWAS p-value | EWAS rank | UCSC RefGene Name | UCSC RefGene Group | Relation to UCSC CpG Island | Regulatory Feature Group | Phantom5 Enhancer | DHS | eQTM |
| --- | --- | --- | --- | --- | --- | --- | --- | --- | --- | --- | --- | --- | --- |
| cg04887278 | 0.6037 | 6 | 83903927 | 3505 | 2.40E-11 | 1 | *PGM3*; *RWDD2A* | TSS1500; 5'UTR | S_Shore |  | 0 | 1 | 0 |
| cg03519157 | 0.4604 | 11 | 46367033 | 3526 | 4.90E-11 | 2 | *DGKZ* | 5'UTR; 1stExon; Body | Island |  | 0 | 1 | 1 |
| cg08469255 | 0.232 | 6 | 30851069 | 3451 | 6.70E-10 | 3 | *DDR1* | TSS1500 | N_Shore | Unclassified | 0 | 1 | 1 |
| cg17810176 | 0.1021 | 19 | 36035831 | 3523 | 1.70E-09 | 4 | *GAPDHS*; *TMEM147* | Body; TSS1500 | N_Shore |  | 0 | 1 | 0 |
| cg22505924 | 0.0791 | 2 | 64488157 | 3527 | 4.10E-09 | 6 |  |  |  | Unclassified | 1 | 1 | 0 |
| cg27496526 | 0.4744 | 15 | 41805530 | 3526 | 6.90E-09 | 7 | *LTK* | Body | Island |  | 0 | 1 | 0 |
| cg25563256 | 0.0349 | 17 | 7341641 | 3524 | 7.60E-09 | 8 | *FGF11* | TSS1500 | N_Shore | Unclassified | 0 | 1 | 0 |
| cg16329896 | 0.1303 | 7 | 47515060 | 3517 | 1.30E-08 | 10 | *TNS3* | 5'UTR |  | Unclassified cell type specific | 1 | 1 | 0 |
| cg10394832 | 0.0612 | 1 | 111889533 | 3525 | 2.50E-08 | 13 | *C1orf88* | Body | Island | Promoter associated | 0 | 0 | 0 |
| cg17496659 | 0.0165 | 1 | 3568245 | 3520 | 3.60E-08 | 16 | *TP73* | TSS1500 | Island |  | 0 | 1 | 0 |
| cg21159993 | 0.019 | 5 | 139554405 | 3523 | 3.70E-08 | 17 | *C5orf32* | TSS1500 | Island | Promoter associated | 0 | 1 | 1 |
| cg19989295 | 0.003 | 14 | 24641077 | 3483 | 5.10E-08 | 19 | *REC8* | TSS200 | Island | Promoter associated | 0 | 1 | 1 |
| cg1382554 | 0.0131 | 9 | 140117257 | 3514 | 6.50E-08 | 21 | *RNF208* | TSS1500 | Island |  | 0 | 1 | 0 |
| cg05302489 | 0.0382 | 6 | 31760426 | 3499 | 1.00E-07 | 28 | *VARS* | Body | N_Shelf |  | 0 | 0 | 1 |
| cg02170695 | -0.0968 | 21 | 38909809 | 3525 | 1.40E-07 | 34 |  |  |  |  | 0 | 0 | 0 |
| cg13067434 | 0.0325 | 15 | 63836865 | 3522 | 1.80E-07 | 37 | *USP3* | Body |  |  | 0 | 0 | 0 |
| cg04382191 | 0.026 | 3 | 9437901 | 3525 | 2.20E-07 | 43 | *SETD5*; *THUMPD3-AS1* | TSS1500; Body | N_Shore | Promoter associated cell type specific | 0 | 1 | 0 |

**Notes:** Chr., chromosome; EWAS, epigenome-wide association study; N\_Shore, north shore; S\_Shore, south shore, UCSC, University of California Santa Cruz Genome Browser; DHS, DNase hypersensitive site; eQTM, expression quantitative trait methylation.

For replication, we chose MESA because it is one of the few comparable aging studies in the US with DNAm data. However, in MESA, DNAm data were profiled on the Illumina Infinium Methylation450 BeadChip. Thus, we also constructed a 450k version of the aSES and cSES biomarkers (aSES-BIO450 and cSES-BIO450) that applied the same parameters and model specifications to CpG sites on the 450k array that overlap with the EPIC array. Elastic net selected 29 of 34,095 sites for aSES (lambda=0.138, CV R^2^=5.34%, *SI Appendix*, Table S4) and 15 of 46,548 sites for cSES (lambda=0.128, CV R^2^=3.28%, *SI Appendix*, Table S5). The aSES-BIO450 included 14 of the same CpG sites as aSES-BIO, and the two biomarkers were highly correlated (r=0.89) (*SI Appendix*, Table S6). The cSES-BIO450 was also highly correlated with cSES-BIO (r=0.99) and shared 13 CpG sites.

### Correlations between SES indices, SES biomarkers, and epigenetic aging measures

In HRS, the measured adult SES (aSES) and childhood SES (cSES) indices were correlated (r=0.43) (*SI Appendix*, Table S6). The aSES index was correlated with aSES-BIO (r=0.24), the cSES index was correlated with cSES-BIO (r=0.19), and the two SES biomarkers were also correlated (r=0.28). aSES-BIO was correlated with both aging clocks (r=0.18 for GrimAge and r=0.50 for DunedinPACE), and cSES-BIO was weakly correlated with GrimAge (r=0.05) and DunedinPACE (r=0.16).

### Associations between SES biomarkers and downstream health outcomes

Figure 2 displays associations between both aSES-BIO and cSES-BIO and downstream health outcomes measured approximately two years after DNAm measurement in the HRS. Both aSES-BIO and cSES-BIO were associated with the number of chronic conditions, the cardiometabolic conditions index (CMCI), self-reported health status (SRHS), and mortality after adjusting for age, sex, race, and smoking (all p<0.0001, Model 1). Associations between aSES-BIO and these same health outcomes persisted in magnitude and statistical significance after adjusting for measured aSES (Model 2). *SI Appendix*, Table S7 shows that these aSES-BIO associations were slightly attenuated but continue to persist after adjusting for the epigenetic aging clocks (Models 3a and 4a) and for both measured aSES and the clocks (Models 4a and 4b). Conversely, associations with cSES-BIO were statistically insignificant after adjusting for both cSES and DunedinPACE (*SI Appendix*, Table S7, Model 4b), and for CMCI, after adjustment for DunedinPACE (Model 3b) or both cSES and GrimAge (Model 4a). Associations between aSES-BIO and Langa-Weir dementia were fully attenuated after adjusting for measured aSES (Models 2, 4a-4b), but persisted after adjusting for the epigenetic aging clocks (Models 3a and 3b). There were no significant associations between cSES-BIO and Langa-Weir dementia. Associations between health outcomes and the aSES-BIO450 or cSES-BIO450 biomarkers were similar to EPIC chip biomarker associations (*SI Appendix*, Table S8).

**Figure 2.**
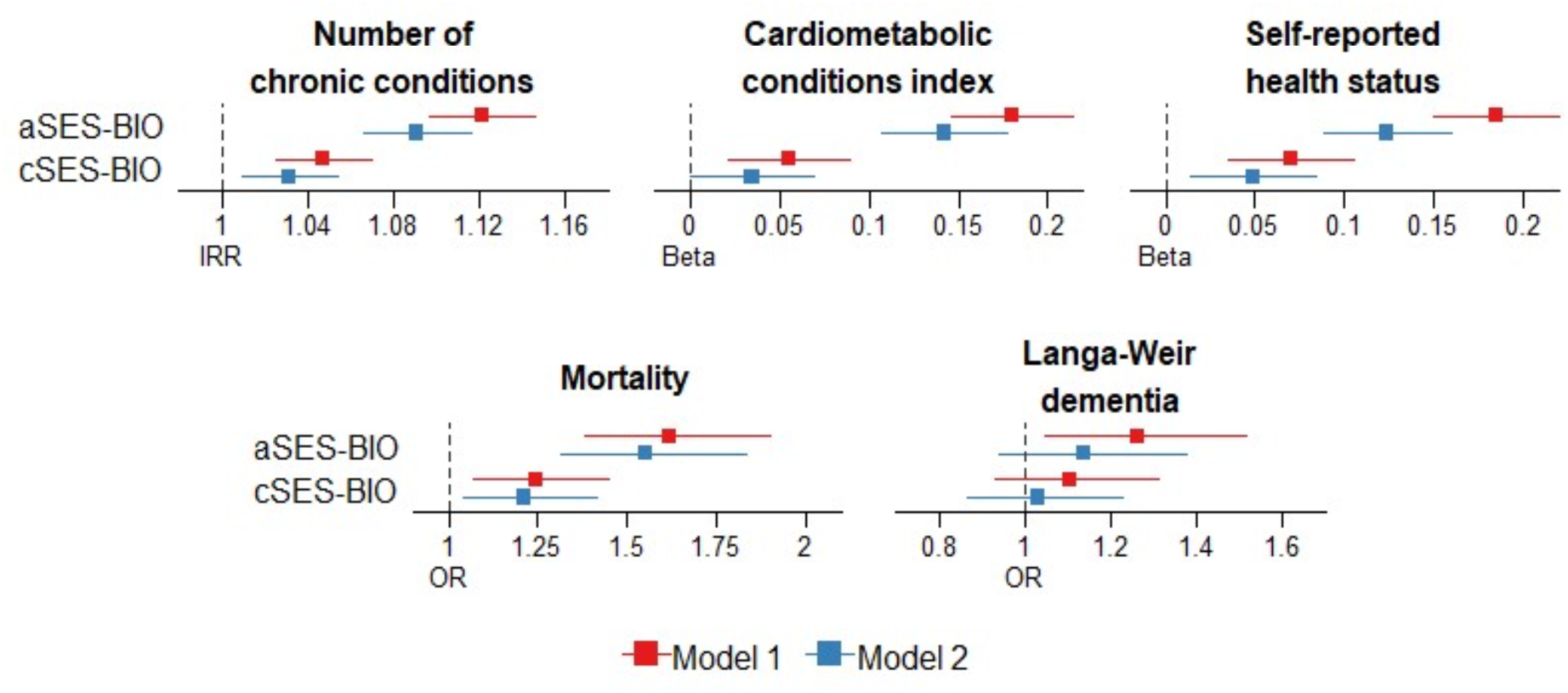
Associations between SES biomarkers and health outcomes in HRS **Notes:** Figure displays estimates from separate regressions of health or mortality on aSES-BIO (adult SES biomarker constructed from EPIC chip methylation sites) or cSES-BIO (childhood SES biomarker constructed from EPIC chip methylation sites). The aSES-BIO and cSES-BIO measures were standardized for analysis. Model 1 controls for age, sex, race, current smoking, former smoking, and *APOE* e4 carrier status (dementia models only). Model 2 adds controls for measured aSES (in aSES-BIO regressions) or cSES (in cSES-BIO regressions). For chronic conditions, a Poisson model was used; for the cardiometabolic conditions index (CMCI) and self-reported health status (SRHS) linear models were used; for mortality and dementia logistic models were used. 95% confidence intervals. IRR, incidence rate ratio; OR, odds ratio. Sample sizes: number of chronic conditions=3120; CMCI=3120; SRHS=3118; mortality=3527; dementia=3120.

To assess the performance of the SES biomarkers relative to measured SES, we also estimated a version of Model 1 (Model 1*) that used the aSES or cSES indices instead of the SES biomarkers (*SI Appendix*, Table S7). In general, SES biomarker associations were not significantly different from aSES or cSES associations in our training sample, except for SRHS and Langa-Weir dementia, where associations were significantly higher in magnitude. Finally, Model 1 associations were robust after adjusting for either measured cSES (in the case of aSES-BIO) or measured aSES (in the case of cSES-BIO) (*SI Appendix*, Table S7, Model 5).

### Replication in MESA

In the replication sample, the adult SES biomarker (aSES-BIO450) was correlated with aSES and explained approximately 4.4% of the variation in aSES (*SI Appendix*, Table S9). Variance explained was similar after accounting for age and sex (partial R^2^=3.88%, p<0.0001). aSES-BIO450 correlations with GrimAge (r=0.36) and DunedinPACE (r=0.60) were higher than correlations in HRS (*SI Appendix*, Table S9). The biomarker also had significant associations with downstream health outcomes that were similar in magnitude to HRS associations, including CMCI, SRHS, and mortality (p<0.05, Models 1, Figure 3), and these associations persisted after controlling for aSES (Model 2). Importantly, aSES-BIO450 appears to be a better predictor of downstream CMCI and mortality than aSES, whereas for SRHS, aSES outperforms the aSES biomarker (*SI Appendix*, Table S10, Model 1* and Model 1). Associations between aSES-BIO450 and SRHS were also fully attenuated after adjusting for GrimAge or DunedinPACE. For CMCI, associations persisted in models that controlled for GrimAge (Models 3a and 4a) but were statistically insignificant after adjusting for DunedinPACE (Models 3b and 4b). Mortality associations persisted across all models but were attenuated in magnitude and significance when adjusting for DunedinPACE. No associations were found between the biomarker and ICD-based all cause dementia.

**Figure 3.**
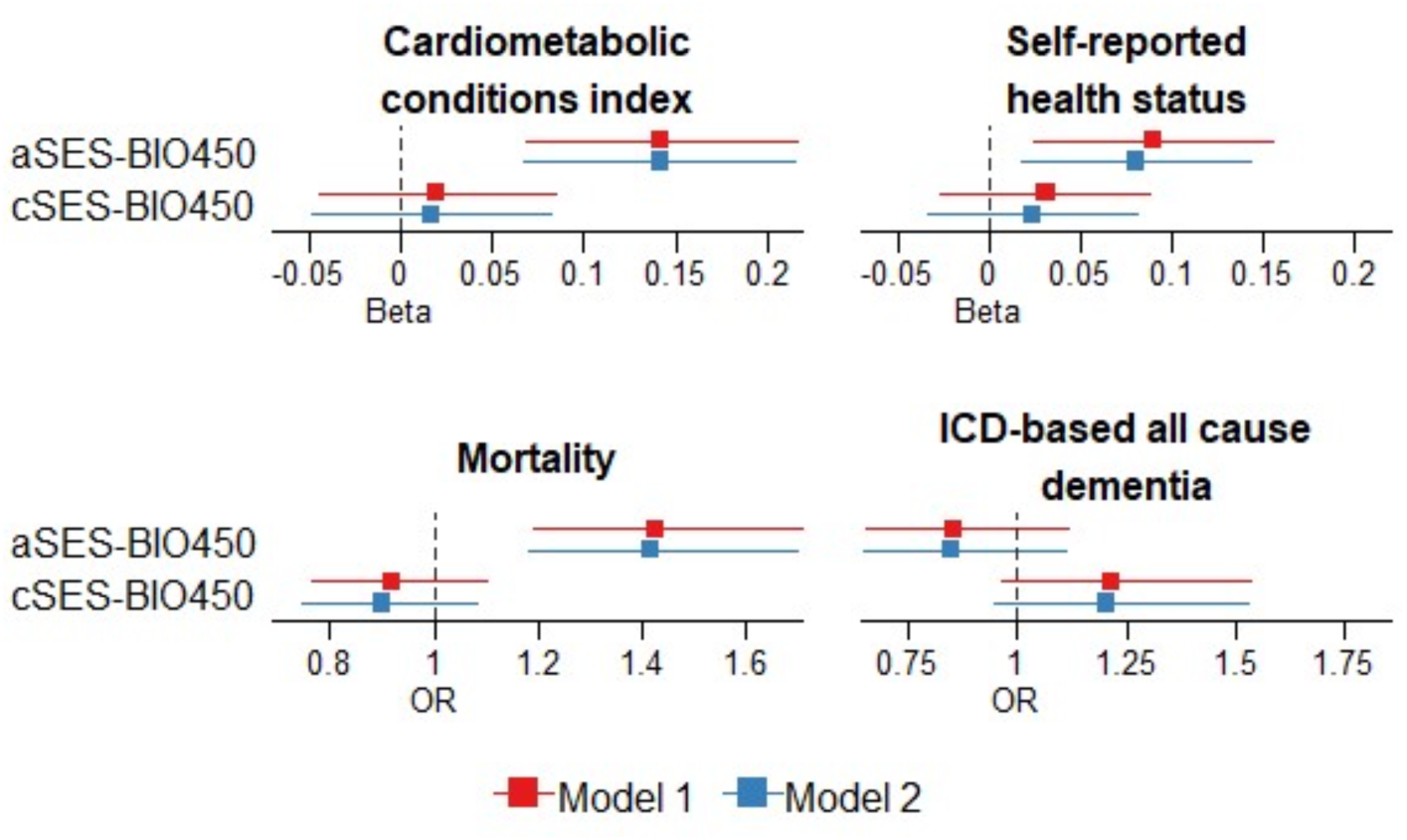
Associations between SES biomarkers and health outcomes in MESA **Notes:** Figure displays estimates from separate regressions of health or mortality on aSES-BIO450 (adult SES biomarker constructed from 450K chip methylation sites) or cSES-BIO450 (childhood SES biomarker constructed from 450k chip methylation sites). The aSES-BIO450 and cSES-BIO450 measures were standardized for analysis. Model 1 controls for age, sex, race, current smoking, former smoking, and *APOE* e4 carrier status (dementia models only). Model 2 adds controls for measured aSES (in aSES-BIO450 regressions) or cSES (in cSES-BIO450 regressions). For the cardiometabolic conditions index (CMCI) and self-reported health status (SRHS) linear models were used; for mortality and dementia logistic models were used. 95% confidence intervals. OR, odds ratio. Sample sizes: CMCI=827; SRHS=848; Mortality=1180; Dementia=1180.

For childhood SES, we were limited to self-reports of parental education and therefore could not test the correlation between cSES-BIO450 and the complete cSES index in MESA. The cSES-BIO450 was marginally correlated with parental education (r=0.07), and the cSES biomarker was not correlated with GrimAge or DunedinPACE (*SI Appendix*, Table S9). Associations between cSES-BIO450 and downstream health outcomes were largely insignificant (*SI Appendix*, Table S10).

### Functional characterization of biomarker CpGs

CpG sites selected for the aSES-BIO were enriched in DNAse hypersensitivity sites (DHS) and enhancer regions, while CpGs included in the cSES-BIO were enriched in promoter regions (*SI Appendix*, Table S11; all p<0.05). Further, compared to CpGs that were not included in the biomarker indices, CpGs selected for both EPIC and 450K SES biomarkers were enriched for eQTM expression (p<8.3E-04) (*SI Appendix*, Table S11). Taken together, these results suggest that the selected CpGs may be important for the regulation of gene expression.

Based on Illumina annotation file mapping, CpGs selected for the aSES-BIO and cSES-BIO mapped to 21 and 18 unique genes, respectively. After FDR correction, gene-set enrichment of GO Biological Processes (BP) terms and KEGG pathways were not observed for either set of CpGs when mapped to their proximal genes (FDR q<0.01). When utilizing eQTM data from Keshawarz et al. (2023), CpGs selected for the aSES-BIO and cSES-BIO mapped to the expression of 230 and 66 genes, respectively, in *cis* or in *trans*. Additionally, CpGs selected for aSES-BIO were associated with the expression of genes enriched for 8 GO BP terms related to cell death, T cell differentiation and activation, cell migration, and cell-cell signaling (*SI Appendix*, Table S12 and Figure S3). CpGs selected for the cSES-BIO were associated with the expression of genes enriched for 13 GO BP terms and 1 KEGG pathway, including chemotaxis, T cell differentiation, immune system process, and cell adhesion molecules (*SI Appendix*, Tables S13 and S14; Figure S4). Results from the GO enrichment analysis for the cSES-BIO450 were similar (*SI Appendix*, Table S15 and Figure S5). There was no observed enrichment of GO or KEGG pathways for genes associated with CpGs selected for the aSES-BIO450.

### Biomarker CpG overlap with published EWAS

Several CpGs selected for both the aSES and cSES biomarkers have been previously associated with diseases and traits identified in large-scale EWAS, including age, chronic diseases, and inflammation. In general, CpG sites in cSES-BIO overlapped more with prior EWAS of immune system processes, whereas aSES-BIO CpGs overlapped with a broader range of traits or molecular processes related to metabolic functioning, health behaviors, or cancer (*SI Appendix*, Tables S16-S19).

## Discussion

We constructed DNAm-based biomarkers of child and adult SES in a large, population-representative sample of older adults (HRS) and tested their performance and association with downstream health outcomes in one of the few comparable samples with DNAm data in the US (MESA). The aSES-BIO explained approximately 4% of the variance in measured aSES in MESA. The explanatory power of cSES-BIO could not be fully ascertained because complete cSES index information used to train the biomarker in HRS were not available in MESA. In both HRS and MESA, aSES-BIO and cSES-BIO were associated with chronic disease, cardiometabolic conditions, and mortality two to six years after DNAm profiling, and these associations persisted after controlling for measured SES. Moreover, although the SES biomarkers were correlated with second-generation epigenetic aging measures, adjusting for GrimAge and DunedinPACE did not fully attenuate these associations. Notably, in our replication sample, the aSES biomarker was a better predictor of downstream cardiometabolic conditions and mortality than measured aSES. Taken together, these findings suggest that DNAm-based surrogates of SES may be useful proxies for unmeasured social exposures and may contain additional valuable information on the biological underpinnings of the SES-health gradient that are not fully captured by self-reported SES or epigenetic aging measures.

Further analyses of the CpG sites that comprise the SES biomarkers revealed enrichment in enhancer and promoter regions, DNase hypersensitivity sites, and eQTMs, suggesting they may be critical for the regulation of gene expression. In gene-set enrichment analysis, selected CpGs were associated with the expression of genes enriched in pathways related to chemotaxis, T cell activation, immune system response, cell migration, and cell death. CpG sites in both biomarkers were associated with prior EWAS of age, age-related chronic diseases, inflammation (e.g., chronic pain, COPD, and C-reactive protein), and clear cell renal carcinoma (56). However, unique patterns for each biomarker also emerged. CpG sites in aSES-BIO were identified in prior EWAS of Type 2 diabetes and health behaviors, including smoking, alcohol use, diet quality, and BMI. Additionally, CpGs were associated with pancreatic cancer (57) and circulating tumor necrosis factor receptor 2 (sTNFR2) (58), a known biomarker for several types of cancer, including colorectal cancer, non-Hodgkin’s lymphoma, and hepatocarcinoma (59), that impacts tumor activation and progression (60). Of note, the top CpG selected for both aSES-BIO and aSESBIO-450K (cg18181703) maps to the body of the suppressor of cytokine signaling 3 (*SOCS3*) gene, which has a role in regulating cytokine and hormone activity. Both inflammation and infection can stimulate the expression of *SOCS3* in different cell populations (61). cg18181703 was also identified in prior EWAS of educational attainment and social disadvantage (62, 63). Finally, although associations between aSES-BIO and dementia were relatively weaker when compared to other downstream health outcomes, one of the CpG sites selected for aSES-BIO (cg06021088) maps to the Bridging Integrator 1 (*BIN1*) gene, which is considered the second most significant genetic risk factor for late-onset Alzheimer’s disease (64, 65).

Conversely, CpGs in the cSES-BIO were found more in prior EWAS of immune and autoimmune diseases, including atopy, Sjorgren’s syndrome, and Rheumatoid arthritis, and a subset of CpGs were associated with the expression of genes enriched for immune system processes and T cell differentiation. These findings support prior research that has shown childhood disadvantage is a risk factor for immune dysregulation (66) and changes in T cell activity (67), which in turn can increase the risk of mental and physical health issues in adulthood, including depression, cardiovascular disease, Type 2 diabetes, and cancer (68, 69). This supports the hypothesis of biological embedding of adult disease in early life, potentially through adverse childhood circumstances that may have cumulative or interactive effects on adult health.

This study has several limitations. First, although we include covariates in our models to mitigate any confounding from sex, race, smoking behavior, genetic ancestry, and technical noise, we cannot draw conclusions regarding the causal relationship between socioeconomic position in childhood or adulthood and DNAm signatures. In addition, self-reported measures of cSES in particular may be subject to recall bias. Thus, the CpG sites chosen by the machine-learning algorithm likely detect a mix of features that are not directly causal, and some may be uninformative correlates of SES. Moreover, some features may be specific to the US context and may not be generalizable to other countries, particularly in other high-income countries where different welfare states or social programs affect the depth and distribution of social disadvantage, and/or in low- and middle-income countries where the biosocial determinants of health may vary due to vastly different socioeconomic and epidemiological contexts (70). Additionally, technical noise from batch effects or unreliable probes may limit generalizability, and SES biomarkers that were trained in the HRS EPIC array data could not be validated in the MESA 450k array data. Further, DNAm was profiled in different tissue types in HRS (whole blood) and MESA (monocytes). Although our results do appear to replicate across these two tissue types, further replication in other cell/tissue samples and across different applications and contexts is needed. Despite these limitations, this study provides a foundation for the rigorous creation of epigenetic markers for social exposures. A major strength of this study is the use of large, multi-racial/ethnic population-representative samples with rich socioeconomic data from both childhood and adulthood. Within both the HRS and MESA samples, we were also able to test longitudinal associations between the aSES and cSES biomarkers and downstream health and mortality. Future analyses include evaluating the SES biomarkers in each racial/ethnic group separately and examining associations between the identified CpG sites and forthcoming RNA-seq data in HRS.

## Materials and Methods

### Data

#### Health and Retirement Study (HRS)

The HRS is a nationally representative, longitudinal panel study of individuals over the age of 50 and their spouses that began in 1992 (71, 72). HRS participants are interviewed every two years about their income and wealth, health and use of health services, work and retirement, and family connections. New cohorts are introduced every six years to offset attrition and death and to maintain a sample size of approximately 20,000 per wave. DNAm data was profiled in 4,103 racially and socioeconomically diverse HRS participants, and 4,018 samples passed quality control (73). Of these, 491 were missing genomic data. A total of 3,527 participants with DNA methylation, genetic, and socioeconomic data were included in the analyses.

Demographic and socioeconomic data were taken from the RAND HRS Longitudinal File 2020 (V1) (74). Race and ethnicity were self-reported. Neighborhood social disadvantage data were taken from the 2006-2016 Psychosocial and Lifestyle Questionnaire and merged to respondents at the Census tract level using restricted data on the location of primary residence from the 2006-2016 waves (75, 76). Neighborhood socioeconomic disadvantage measures were constructed using the restricted HRS Contextual Data Resource, which contains data from the 2005-2018 American Community Survey linked to respondents at the Census tract level (77). Restricted three-digit Census occupation codes from the 1992-2016 (78) waves were used to merge occupational prestige measures from the General Social Survey (79).

#### DNA methylation (DNAm)

DNAm was measured from whole blood samples collected in the 2016 HRS Venous Blood Study (80). Methylation was measured using the Illumina Infinium Methylation EPIC BeadChip. The *minfi* package in R software was used for data preprocessing and quality control. Samples with mismatched sex or an average median intensity <8.5 were removed. A detection p-value <0.01 was used to remove probes or samples with >5% missing data. Cross-reactive probes were removed prior to elastic net analyses (81). After quality control, a total of 836,660 CpGs were available for analysis. DNA methylation at each CpG site was quantified using beta values, which approximate the proportion of methylation. The Houseman method was used to estimate white blood cell (WBC) proportions(82). We also used two epigenetic aging measures, GrimAge (83) and Dunedin Pace of Aging (DunedinPACE) (25). GrimAge was calculated and released by HRS (73), and we calculated PACE using the *DunedinPACE* R package (25).

#### Genotypes

We used genotype data to estimate genetic principal components (PCs) of ancestry and to obtain *APOE ε*4 carrier status. Genotype data for over 18,000 HRS participants was measured using Illumina HumanOmni2.5 BeadChips (HumanOmni2.5-4v1, HumanOmni2.5-8v1) (84). Individuals with call rates <98%, SNPs with call rates <98%, HWE p-value<0.0001, chromosomal anomalies, and first-degree relatives in the HRS were removed. Principal components were calculated using the *SNPRelate* package in R (85). *APOE* isoforms were inferred from SNP genotypes determined by TaqMan assays or imputed using the HumanOmni2.5 data (86). People with at least one copy of the *ε*4 allele were considered *ε*4 carriers.

#### Socioeconomic status

As in our previous work (33), to capture the multi-dimensional nature of SES we used six measures to construct an aSES index: educational attainment, household income, wealth, occupational prestige, neighborhood socioeconomic disadvantage, and neighborhood social environment. Additional details on SES index construction are provided in *SI Appendix*, Section 1. Briefly, we took the unweighted average of the non-missing quintiles for each indicator and reverse coded the indicators so that a higher quintile corresponds to worse SES. <u>Educational attainment</u> was measured in years and was coded as 1=>16 years, 2=16 years, 3=13-15 years, 4=12 years, and 5=<12 years. Similarly, quintiles were constructed after averaging values across all HRS waves up to 2016 for <u>household income</u> (inflation adjusted to 2016 dollars), <u>wealth</u> (total assets inflation adjusted to 2016 dollars), and <u>occupational disadvantage</u> (87) (attained by matching occupational prestige codes from the General Social Survey to respondent three-digit occupation codes). Quintiles for the <u>neighborhood socioeconomic disadvantage score</u> (a summary of six census tract-level metrics reflecting education, occupation, income/wealth, poverty, employment, and housing) and <u>neighborhood social disadvantage score</u> (a summary of conditional Bayes’ estimates for aggregate-level responses on aesthetic quality, safety, and social cohesion) were averaged across 2012-2016 waves and 2006-2016 waves, respectively, to reflect cumulative neighborhood exposures (88, 89). Respondents were required to have non-missing values for at least three of the six measures to be assigned an aSES index. As a robustness check, we also used principal components (PC) analysis of these six variables to construct a PC-based SES measure, which was highly correlated with the SES index (r=0.97). We opted to use the method above to construct the SES index because the PC method required all score elements to be non-missing.

We created a cSES index by taking the unweighted average of two measures: parental education and childhood financial strain (90). Parental education was the average of maternal and paternal education, coded as 0=16 years or more, or 1=13-15 years, 2=12 years, 3=8-11 years, 4=<8 years. For missing values of maternal or paternal education, we first imputed continuous education years from demographic and SES variables using a multivariate, regression-based procedure in Imputation and Variance Estimation (IVEware) software (http://www.isr.umich.edu/src/smp/ive/) as described in Faul, et al. (90). For HRS participants from the earliest wave of the study, parental education was collected as a dichotomous variable; for these participants, we used parental education imputations from Vable, et al. (91) rounded to the nearest year. Childhood financial strain was the sum of whether the respondent has ever moved as a child, received financial help from relatives, had a period of paternal unemployment, or had poor/varied financial status.

#### Health outcomes

To assess health and aging in HRS, we used data from the 2018 wave, which was collected two years after DNAm measurement. Number of chronic conditions (range 0-8) was a count of the self-reported number of conditions that the respondent had ever experienced, including hypertension, diabetes, cancer, lung disease, heart problems, stroke, psychiatric problems, and arthritis. The cardiometabolic conditions index (CMCI) was the mean standardized sum of four mean standardized doctor-diagnosed chronic diseases (diabetes, heart problems, stroke, and hypertension). Self-reported health status (SRHS, range 1-5) was the respondent rating of their overall health, ranging from 1=excellent to 5=poor. Mortality was an indicator (1=yes) of whether the respondent died between the 2016 and 2018 data collection waves. Dementia was evaluated using the Langa-Weir definition (92, 93), which sets dementia classification (1=yes) if an individual scores 0-6 points on a 27-point cognition scale.

#### Replication sample: Multi-Ethnic Study of Atherosclerosis (MESA)

We evaluated the performance of our SES biomarkers in MESA. DNAm was measured from monocytes at Exam 5 (2010–2012) using the Illumina Infinium Methylation450 BeadChip. See Liu et al. (2013) for a description of data processing (94). Briefly, probes were excluded if they had ‘detected’ methylation levels in <90% of MESA samples (detection p-value threshold=0.05), overlapped with a non-unique genomic region, or were designed to assay polymorphic single nucleotide polymorphisms (SNPs) rather than methylation (95). Genomic principal components were estimated with the *EIGENSTRAT* program using genotype data from the Affymetrix 6.0 SNP array (96). *APOE* genotypes were estimated from imputed genotype data and individuals with at least one copy of the *ε*4 allele were considered *ε*4 carriers. Race and ethnicity were self-reported.

The same six SES measures (educational attainment, household income, wealth, occupational disadvantage, neighborhood socioeconomic disadvantage, and neighborhood social environment) were used to construct the adult SES index in MESA. These were coded similarly to HRS except for income and wealth, both of which were not measured continuously (see *SI Appendix, Section 1* for additional details). For income, respondents selected their household’s income bracket from a series of unfolding brackets rather than reporting a continuous value. For wealth, a combination of home and land ownership, car ownership, and investments were used to construct a 5-level categorical variable to take the place of the quintiles in the aSES index (1=home or land + car + investments; 2=home or land + car or investments; 3=home or land only; 4=car or investments; 5=none). In MESA, measures of childhood SES were limited to parental education. Here, we took the average of maternal and paternal education, which was coded as a categorical variable (4=no schooling; 3=some schooling but did not complete HS; 2=HS degree, 1=some college, 0=college/graduate professional degree).

Health outcomes were constructed in the same manner as HRS, except for the number of chronic diseases, which was not available in MESA, and dementia, which was assessed using ICD-based all cause dementia diagnoses through 2018 from the MESA dementia events file. Apart from dementia, all health outcomes were measured at Exam 6, which occurred approximately six years after DNAm data collection (2016–2019).

### Statistical Methods

#### Epigenome-wide association studies (EWAS)

Due to the large number of CpG sites present on the EPIC chip, we used a prescreening procedure to reduce the number of CpGs evaluated when constructing the SES biomarkers by first conducting EWAS for aSES and cSES, separately, in the training data (HRS). EWAS CpGs with p<0.05 were then carried forward into the biomarker construction stage (Figure 1). Our EWAS model was as follows:

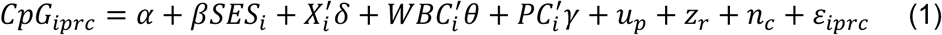

where *SES_i_* is either the aSES or cSES index for individual *i*. The matrix *X_i_* contains individual characteristics including age, sex, and race; due to the strong effects of smoking on the methylome and the well-established socioeconomic gradient in smoking, we also include dichotomous indicators for current and/or former smoking (omitted category=never smoker). Because DNAm was profiled in whole blood and methylation patterns differ by blood cell type, we accounted for differences in white blood cell (*WBC_i_*) composition across samples, or the proportion of monocytes, natural killer cells, B-cells, CD4 T-cells, and CD8 T-cells (omitted category=proportion granulocytes). The first four genetic ancestry PCs were included to account for potential confounding due to ancestry (*PC_i_*). Technical covariates for plate (*p*), plate row (*r*), and plate column (*c*) were included as random effects to account for potential batch effects, or *u_p_*, *z_r_*, and *n_c_*, respectively. Manhattan plots for these EWAS can be found in (*SI Appendix*, Figures S1-S2).

To examine which of the SES index components had the strongest associations with the CpG sites selected into the SES index biomarkers, we also conducted separate EWAS for each of the components of the aSES and cSES indices (Figure 1). We used the same model as above except that education years was included as a covariate when neighborhood socioeconomic disadvantage or neighborhood social environment was the SES measure of interest.

#### SES index biomarker construction

To minimize overfitting and select the CpG sites that are most predictive of the SES indices, we used elastic net, a regression method that performs variable selection and regularization (shrinkage) simultaneously (97). For aSES and cSES separately, we first selected all CpG sites with p<0.05 from the corresponding EWAS, and then adjusted these CpGs for all covariates in the EWAS model. Next, we used the residual CpGs as input for elastic net, using the following parameters in the *glmnet* package in R: alpha=0.5 (equally balancing ridge regression and lasso) and 5-fold cross validation to find the optimal value of lambda (penalty strength parameter) for minimizing prediction error (98). To construct the SES biomarker for adult SES (aSES-BIO) and childhood SES (cSES-BIO), we summed the methylation value at each CpG site selected by elastic net at the optimal value of lambda, weighted by its corresponding beta coefficient, and then standardized the sums. We then calculated the correlation between the methylation-based biomarkers (aSES-BIO and cSES-BIO), measured markers of SES (aSES and cSES), and the epigenetic aging measures (GrimAge and DunedinPACE).

We recognize that many studies, including our replication sample (MESA), have DNAm measured using the Illumina 450K chip rather than the EPIC chip. Therefore, we also constructed a version of the aSES and cSES biomarkers by first restricting to only those EPIC CpG sites that were also included on the 450K chip and then used the same parameters and model specifications for elastic net. We assessed the level of CpG overlap and correlation between these 450K SES biomarkers (aSES-BIO450 and cSES-BIO450) and the EPIC chip biomarkers (aSES-BIO and cSES-BIO).

#### Associations between SES biomarkers and downstream health outcomes

We used Poisson regression (number of chronic conditions), linear regression (CMCI, SRHS), or logistic regression (mortality, dementia) to assess the relationship between the SES biomarkers (aSES-BIO, cSES-BIO) and downstream health and mortality in 2018, or two years after DNAm was profiled in the HRS, using the following model (Model 1):

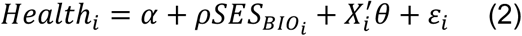

Where *SES_BIO_*_’_ is the aSES-BIO or cSES-BIO biomarker for individual *i* and the matrix *X_i_* includes controls for age, sex, race, and smoking status as in Equation 1. The aSES, cSES, aSES-BIO, and cSES-BIO measures were standardized to have a mean of 0 and standard deviation (SD) of 1 for analysis. For dementia models, we also included an indicator for APOE e4 status. We then examined associations between the SES biomarkers and health outcomes after accounting for the measured SES index (aSES or cSES; Model 2), GrimAge or DunedinPACE (Models 3a and 3b), both measured SES and GrimAge or DunedinPACE (Models 4a and 4b), and either measured cSES (for aSES-BIO) or aSES (for cSES-BIO) to examine SES-BIO associations after accounting for SES in childhood or adulthood (Model 5). To assess the performance of the SES biomarkers relative to measured SES, we also estimated a version of Model 1 (Model 1*) that used the aSES or cSES indices instead of the SES biomarkers. Coefficient p-values<0.05 were considered statistically significant.

#### Replication in MESA

For each MESA participant, aSES-BIO450 or cSES-BIO450 was constructed by multiplying the elastic net beta coefficients by the methylation beta values at the corresponding CpG sites (measured in Exam 5) and taking the standardized sum. We then tested the association of aSES-BIO450 or cSES-BIO450 with measured aSES or cSES and with health outcomes (CMCI, SRHS, mortality, and dementia) collected at MESA Exam 6, which was conducted approximately 6 years after DNAm measurement, or between 2016 and 2019. Number of chronic conditions was not analyzed in MESA, as the data available in MESA did not allow us to create this variable in a comparable way to HRS.

#### Functional characterization of biomarker CpGs

##### Genomic feature enrichment analysis

We performed genomic feature enrichment analysis on the sets of CpG sites selected for each of the SES biomarkers separately. For each set of CpGs, we examined whether their genomic locations were enriched for features including gene promoters, enhancers, DNase I hypersensitivity sites (DHS), CpG islands (CGI), and CpG island flanking shores or shelves (<4 kb from CGI). We used annotation files from Illumina and the UCSC genome browser to identify target genes and genomic features associated with each CpG (99, 100). A CpG site was in the promoter region if it was located 0-1500 bases upstream of a transcription start site. To identify CpGs associated with gene expression, we used results from expression quantitative trait methylation (eQTM) analysis performed using the 450k chip (101) and the EPIC (102) in the Framingham Heart Study (FHS). For each set of CpGs selected for a given biomarker, we compared whether the set of CpGs were more likely to be eQTMs than CpGs that were not selected by comparing the proportion of eQTMs in the two groups. All enrichment analyses were conducted using a two-sided Fisher’s exact test with a significance threshold of p-value<0.05.

##### Gene-set analysis

To better understand the functional pathways of the CpGs selected for each SES biomarker, we conducted Gene Ontology (GO) and Kyoto Encyclopedia of Genes and Genomes (KEGG) pathway analyses using two distinct methods. First, we performed a gene-set analysis using GOmeth from the *missMethyl* package in R (103). This method accounts for potential sources of bias, including the uneven distribution of probes across the epigenome and the lack of independence between CpGs and their associated genes. However, a major limitation of this method is that it uses the chromosomal position of the CpGs to map them to proximal genes. This may not be optimal because CpG sites do not always act on the nearest gene, but instead may influence expression of distal genes. As a second method, we performed GO and KEGG enrichment analysis using the *clusterProfiler* package in R to identify enrichment of biological processes (104). This method allows us to utilize methylation-gene expression pairs (eQTMs) to define the gene list, which places emphasis on the potential functional role of the CpG. For each SES biomarker, our gene signal list was comprised of all genes from an FHS-identified eQTM that mapped to any of the CpGs included in the biomarker. The background gene list included all genes that were part of an eQTM that overlapped with CpGs that were present in our study. FDR-q<0.05 was considered significant for all GO and KEGG pathway analyses.

##### CpG associations in published EWAS

We examined whether each biomarker CpG had been previously associated with any diseases or traits using The EWAS Catalog (105). For each CpG, all traits with p<0.0001 are reported.

## Data Availability

This study used restricted individual level information from the HRS, and our contractual agreement does not permit public dissemination of the data. Details on how to access restricted data for the HRS can be found at https://hrs.isr.umich.edu/data-products/restricted-data. Data used in this analysis from MESA can be obtained through the MESA Data Coordinating Center (https://www.mesanhlbi.org/) and through the database of Genotypes and Phenotypes (dbGaP; phs000209). Analysis code is posted on github: https://github.com/laurenschmitz/epigenetic-SES-biomarker.

## Acknowledgements

The authors thank the other investigators, the staff, and the participants of the HRS and MESA studies for their valuable contributions.

## References

1. A. Danese, B. S. McEwen, Adverse childhood experiences, allostasis, allostatic load, and age-related disease. Physiol Behav 106, 29–39 (2012).

2. S. W. Cole, Social Regulation of Human Gene Expression: Mechanisms and Implications for Public Health. Am J Public Health 103, S84 (2013).

3. L. L. Schmitz, et al., The Socioeconomic Gradient in Epigenetic Ageing Clocks: Evidence from the Multi-Ethnic Study of Atherosclerosis and the Health and Retirement Study. Epigenetics 17, 589–611 (2022).

4. L. L. Schmitz, et al., Associations of Early-Life Adversity With Later-Life Epigenetic Aging Profiles in the Multi-Ethnic Study of Atherosclerosis. Am J Epidemiol 192, 1991–2005 (2023).

5. J. Cerutti, A. A. Lussier, Y. Zhu, J. Liu, E. C. Dunn, Associations between indicators of socioeconomic position and DNA methylation: a scoping review. Clin Epigenetics 13 (2021).

6. H. M. Byun, et al., Temporal Stability of Epigenetic Markers: Sequence Characteristics and Predictors of Short-Term DNA Methylation Variations. PLoS One 7, e39220 (2012).

7. L. Shang, et al., meQTL mapping in the GENOA study reveals genetic determinants of DNA methylation in African Americans. Nature Communications 2023 14:1 14, 1–16 (2023).

8. C. Ladd-Acosta, M. D. Fallin, The role of epigenetics in genetic and environmental epidemiology. Epigenomics [Preprint] (2016).

9. C. Ladd-Acosta, M. D. Fallin, DNA Methylation Signatures as Biomarkers of Prior Environmental Exposures. Current Epidemiology Reports 2019 6:1 6, 1–13 (2019).

10. C. Ladd-Acosta, Epigenetic Signatures as Biomarkers of Exposure. Curr Environ Health Rep 2, 117–125 (2015).

11. C. Ladd-Acosta, M. D. Fallin, The role of epigenetics in genetic and environmental epidemiology. Epigenomics 8, 271–283 (2016).

12. C. Ladd-Acosta, M. D. Fallin, DNA Methylation Signatures as Biomarkers of Prior Environmental Exposures. Current Epidemiology Reports 2019 6:1 6, 1–13 (2019).

13. S. Horvath, DNA methylation age of human tissues and cell types. Genome Biol 14, 1–20 (2013).

14. S. Bocklandt, et al., Epigenetic Predictor of Age. PLoS One 6, e14821 (2011).

15. S. Horvath, et al., Epigenetic clock for skin and blood cells applied to Hutchinson Gilford Progeria Syndrome and ex vivo studies. Aging (Albany NY) 10, 1758 (2018).

16. P. Garagnani, et al., Methylation of ELOVL2 gene as a new epigenetic marker of age. Aging Cell 11, 1132–1134 (2012).

17. G. Hannum, et al., Molecular Cell Resource Genome-wide Methylation Profiles Reveal Quantitative Views of Human Aging Rates. Mol Cell 49, 359–367 (2013).

18. I. Florath, K. Butterbach, H. Müller, M. Bewerunge-hudler, H. Brenner, Cross-sectional and longitudinal changes in DNA methylation with age: an epigenome-wide analysis revealing over 60 novel age-associated CpG sites. Hum Mol Genet 23, 1186–1201 (2014).

19. S. Horvath, et al., Aging effects on DNA methylation modules in human brain and blood tissue. Genome Biol 13, R97 (2012).

20. C. I. Weidner, et al., Aging of blood can be tracked by DNA methylation changes at just three CpG sites. Genome Biol 15, 1–12 (2014).

21. Y. Zhang, et al., DNA methylation signatures in peripheral blood strongly predict all-cause mortality. Nature Communications 2017 8:1 8, 1–11 (2017).

22. M. E. Levine, et al., An epigenetic biomarker of aging for lifespan and healthspan. Aging (Albany NY) 10, 573 (2018).

23. A. T. Lu, et al., DNA methylation GrimAge strongly predicts lifespan and healthspan. Aging (Albany NY) 11, 303 (2019).

24. D. W. Belsky, et al., Quantification of the pace of biological aging in humans through a blood test, the DunedinPoAm DNA methylation algorithm. Elife 9, 1–56 (2020).

25. D. W. Belsky, et al., DunedinPACE, A DNA methylation biomarker of the Pace of Aging. Elife 11 (2022).

26. R. Duan, Q. Fu, Y. Sun, Q. Li, Epigenetic clock: A promising biomarker and practical tool in aging. Ageing Res Rev 81, 101743 (2022).

27. J. D. Faul, et al., Epigenetic-based age acceleration in a representative sample of older Americans: Associations with aging-related morbidity and mortality. Proc Natl Acad Sci U S A 120, e2215840120 (2023).

28. M. E. Levine, A. T. Lu, D. A. Bennett, S. Horvath, Epigenetic age of the pre-frontal cortex is associated with neuritic plaques, amyloid load, and Alzheimer’s disease related cognitive functioning. Aging (Albany NY) 7, 1198 (2015).

29. L. Perna, et al., Epigenetic age acceleration predicts cancer, cardiovascular, and all-cause mortality in a German case cohort. Clin Epigenetics 8, 1–7 (2016).

30. R. E. Marioni, et al., DNA methylation age of blood predicts all-cause mortality in later life. Genome Biol 16, 1–12 (2015).

31. E. M. Crimmins, B. Thyagarajan, M. E. Levine, D. R. Weir, J. Faul, Associations of Age, Sex, Race/Ethnicity, and Education With 13 Epigenetic Clocks in a Nationally Representative U.S. Sample: The Health and Retirement Study. The Journals of Gerontology: Series A 76, 1117–1123 (2021).

32. L. Raffington, et al., Socioeconomic disadvantage and the pace of biological aging in children. Pediatrics 147 (2021).

33. L. L. Schmitz, et al., The Socioeconomic Gradient in Epigenetic Ageing Clocks: Evidence from the Multi-Ethnic Study of Atherosclerosis and the Health and Retirement Study. Epigenetics 17, 589–611 (2022).

34. D. L. McCartney, et al., Epigenetic prediction of complex traits and death. Genome Biol 19, 136 (2018).

35. R. F. Hillary, R. E. Marioni, G. Sharp, MethylDetectR: a software for methylation-based health profiling [version 1; peer review: 1 approved with reservations] report. (2020). 10.12688/wellcomeopenres.16458.1.

36. A. Cappozzo, et al., A blood DNA methylation biomarker for predicting short-term risk of cardiovascular events. Clin Epigenetics 14, 1–17 (2022).

37. D. A. Gadd, et al., Epigenetic scores for the circulating proteome as tools for disease prediction. Elife 11 (2022).

38. Y. Zhang, et al., Smoking-associated DNA methylation markers predict lung cancer incidence. Clin Epigenetics 8, 1–12 (2016).

39. R. M. Walker, et al., Assessment of dried blood spots for DNA methylation profiling. Wellcome Open Res 4 (2019).

40. R. Joehanes, et al., Epigenetic Signatures of Cigarette Smoking. Circ Cardiovasc Genet 9, 436–447 (2016).

41. R. C. Richmond, M. Suderman, R. Langdon, C. L. Relton, G. D. Smith, DNA methylation as a marker for prenatal smoke exposure in adults. Int J Epidemiol 47, 1120 (2018).

42. E. Colicino, et al., Blood DNA methylation biomarkers of cumulative lead exposure in adults. J Expo Sci Environ Epidemiol 31, 108–116 (2021).

43. F. Guida, et al., Dynamics of smoking-induced genome-wide methylation changes with time since smoking cessation. Hum Mol Genet 24, 2349–2359 (2015).

44. M. Moqri, et al., Leading Edge Biomarkers of aging for the identification and evaluation of longevity interventions. (2023). 10.1016/j.cell.2023.08.003.

45. N. Krieger, A glossary for social epidemiology. J Epidemiol Community Health (1978) 55, 693–700 (2001).

46. E. M. Crimmins, Social hallmarks of aging: Suggestions for geroscience research. Ageing Res Rev 63, 101136 (2020).

47. B. G. Link, J. Phelan, Social conditions as fundamental causes of disease. J Health Soc Behav Spec No, 80–94 (1995).

48. K. A. Muscatell, S. N. Brosso, K. L. Humphreys, Socioeconomic status and inflammation: a meta-analysis. Molecular Psychiatry 2018 25:9 25, 2189–2199 (2018).

49. S. J. Lupien, B. S. McEwen, M. R. Gunnar, C. Heim, Effects of stress throughout the lifespan on the brain, behaviour and cognition. Nature Reviews Neuroscience 2009 10:6 10, 434–445 (2009).

50. B. L. Needham, et al., Life course socioeconomic status and DNA methylation in genes related to stress reactivity and inflammation: The multi-ethnic study of atherosclerosis. Epigenetics 10, 958–969 (2015).

51. C. Power, C. Hertzmant, Social and biological pathways linking early life and adult disease. Br Med Bull 53, 210–221 (1997).

52. B. Galobardes, G. D. Smith, J. W. Lynch, Systematic Review of the Influence of Childhood Socioeconomic Circumstances on Risk for Cardiovascular Disease in Adulthood. Ann Epidemiol 16, 91–104 (2006).

53. L. A. Gavrilov, N. S. Gavrilova, Early-Life Programming of Aging and Longevity: The Idea of High Initial Damage Load (the HIDL Hypothesis). Ann N Y Acad Sci 1019, 496–501 (2004).

54. D. J. P. Barker, The fetal and infant origins of adult disease. BMJ : British Medical Journal 301, 1111 (1990).

55. A. Hüls, D. Czamara, Methodological challenges in constructing DNA methylation risk scores. Epigenetics 15, 1–11 (2020).

56. M. B. Wozniak, et al., Integrative Genome-Wide Gene Expression Profiling of Clear Cell Renal Cell Carcinoma in Czech Republic and in the United States. PLoS One 8, e57886 (2013).

57. K. Nones, et al., Genome-wide DNA methylation patterns in pancreatic ductal adenocarcinoma reveal epigenetic deregulation of SLIT-ROBO, ITGA2 and MET signaling. Int J Cancer 135, 1110–1118 (2014).

58. M. M. Mendelson, et al., Epigenome-wide association study of soluble tumor necrosis factor receptor 2 levels in the framingham heart study. Front Pharmacol 9 (2018).

59. A. E. R. Kartikasari, et al., Elevation of circulating TNF receptor 2 in cancer: A systematic meta-analysis for its potential as a diagnostic cancer biomarker. Front Immunol 13, 918254 (2022).

60. Y. Sheng, F. Li, Z. Qin, TNF receptor 2 makes tumor necrosis factor a friend of tumors. Front Immunol 9, 370323 (2018).

61. B. Carow, M. E. Rottenberg, SOCS3, a Major Regulator of Infection and Inflammation. Front Immunol 5 (2014).

62. Y. Z. Wang, et al., DNA Methylation Mediates the Association Between Individual and Neighborhood Social Disadvantage and Cardiovascular Risk Factors. Front Cardiovasc Med 9, 848768 (2022).

63. J. van Dongen, et al., DNA methylation signatures of educational attainment. NPJ Sci Learn 3, 7 (2018).

64. A. Sudwarts, et al., BIN1 is a key regulator of proinflammatory and neurodegeneration-related activation in microglia. Mol Neurodegener 17, 33 (2022).

65. O. Saha, et al., The Alzheimer’s disease risk gene BIN1 regulates activity-dependent gene expression in human-induced glutamatergic neurons. Mol Psychiatry 1–13 (2024).

66. S. Ravi, M. J. Shanahan, B. Levitt, K. M. Harris, S. W. Cole, Socioeconomic inequalities in early adulthood disrupt the immune transcriptomic landscape via upstream regulators. Scientific Reports 2024 14:1 14, 1–11 (2024).

67. M. M. C. Elwenspoek, et al., Proinflammatory T Cell Status Associated with Early Life Adversity. The Journal of Immunology 199, 4046–4055 (2017).

68. M. A. Chen, et al., Immune and Epigenetic Pathways Linking Childhood Adversity and Health Across the Lifespan. Front Psychol 12 (2021).

69. K. M. Ziol-Guest, G. J. Duncan, A. Kalil, W. T. Boyce, Early childhood poverty, immune-mediated disease processes, and adult productivity. Proc Natl Acad Sci U S A 109, 17289–17293 (2012).

70. T. W. McDade, Three common assumptions about inflammation, aging, and health that are probably wrong. Proceedings of the National Academy of Sciences 120, e2317232120 (2023).

71. F. T. Juster, R. Suzman, An Overview of the Health and Retirement Study. J Hum Resour 30, S7 (1995).

72. A. Sonnega, et al., Cohort Profile: the Health and Retirement Study (HRS). Int J Epidemiol 43, 576–585 (2014).

73. E. Crimmins, J. K. Kim, HRS Documentation Report HRS Epigenetic Clocks. (2020).

74. [dataset] RAND HRS Longitudinal File 2020. Produced by the RAND Center for the Study of Aging, with funding from the National Institute on Aging and the Social Security Administration. Santa Monica, CA (March 2023).

75. [dataset] Smith, J., Ryan, L. H., Fisher, G. G., Sonnega, A., & Weir, D. R. (2017). HRS psychosocial and lifestyle questionnaire 2006–2016. Survey Research Center, Institute for Social Research, University of Michigan.

76. [dataset] Health and Retirement Study, Restricted Cross-Wave Geographic Information (Detail) [1992-2020]. Survey Research Center, Institute for Social Research, University of Michigan.

77. [dataset] Ailshire J, Mawhorter S, Choi EY. Contextual Data Resource (CDR): United States Decennial Census and American Community Survey Data, 1990–2018, Version 2.0. Los Angeles, CA: USC/UCLA Center on Biodemography and Population Health. (2020).

78. [dataset] Health and Retirement Study, Restricted Industry and Occupation Data [1992-2020]. Survey Research Center, Institute for Social Research, University of Michigan.

79. [dataset] Smith, TW and Son, J. The General Social Survey, Occupational Prestige Ratings. NORC, University of Chicago. (2014).

80. Venous Blood Collection and Assay Protocol in the 2016 Health and Retirement Study | Health and Retirement Study. Available at: https://hrs.isr.umich.edu/publications/biblio/9065 [Accessed 4 April 2024].

81. R. Pidsley, et al., Critical evaluation of the Illumina MethylationEPIC BeadChip microarray for whole-genome DNA methylation profiling. Genome Biol 17, 1–17 (2016).

82. E. A. Houseman, et al., DNA methylation arrays as surrogate measures of cell mixture distribution. BMC Bioinformatics 13, 1–16 (2012).

83. A. T. Lu, et al., DNA methylation GrimAge strongly predicts lifespan and healthspan. Aging 11, 303–327 (2019).

84. [dataset] Health and Retirement Study, Genetic Data. Produced and distributed by the University of Michigan with Funding from the National Institute on Aging (grant number NIA U01AG009740) Ann Arbor, MI (2021).

85. X. Zheng, et al., A high-performance computing toolset for relatedness and principal component analysis of SNP data. Bioinformatics 28, 3326–3328 (2012).

86. [dataset] J.D. Faul, et al., Health and Retirement Study: APOE and Serotonin Transporter Alleles – Early Release. (2021).

87. [dataset] Hout M, Smith TW, Marsden PV. Prestige and Socioeconomic Scores for the 2010 Census Codes. (2016).

88. A. V. D. Roux, et al., Neighborhood of Residence and Incidence of Coronary Heart Disease. New England Journal of Medicine 345, 99–106 (2001).

89. M. S. Mujahid, A. V. Diez Roux, J. D. Morenoff, T. Raghunathan, Assessing the Measurement Properties of Neighborhood Scales: From Psychometrics to Ecometrics. Am J Epidemiol 165, 858–867 (2007).

90. J. D. Faul, et al., Trans-Ethnic Meta-Analysis of Interactions Between Genetics and Early-Life Socioeconomic Context on Memory Performance and Decline in Older Americans. J Gerontol A Biol Sci Med Sci 77, 2248–2256 (2022).

91. A. M. Vable, P. Gilsanz, T. T. Nguyen, I. Kawachi, M. M. Glymour, Validation of a theoretically motivated approach to measuring childhood socioeconomic circumstances in the Health and Retirement Study. PLoS One 12 (2017).

92. E. M. Crimmins, J. K. Kim, K. M. Langa, D. R. Weir, Assessment of Cognition Using Surveys and Neuropsychological Assessment: The Health and Retirement Study and the Aging, Demographics, and Memory Study. The Journals of Gerontology: Series B 66B, i162–i171 (2011).

93. K.M. Langa, D.R. Weir, M. Kabeto, A. Sonnega, Langa-Weir Classification of Cognitive Function (1995-2020). Survey Research Center, Institute for Social Research, University of Michigan. (2023).

94. Y. Liu, et al., Methylomics of gene expression in human monocytes. Hum Mol Genet 22, 5065–5074 (2013).

95. R. Pidsley, et al., A data-driven approach to preprocessing Illumina 450K methylation array data. BMC Genomics 14, 1–10 (2013).

96. A. L. Price, et al., Principal components analysis corrects for stratification in genome-wide association studies. Nature Genetics 2006 38:8 38, 904–909 (2006).

97. H. Zou, T. Hastie, Regularization and Variable Selection Via the Elastic Net. J R Stat Soc Series B Stat Methodol 67, 301–320 (2005).

98. J. Friedman, T. Hastie, R. Tibshirani, Regularization Paths for Generalized Linear Models via Coordinate Descent. J Stat Softw 33, 1 (2010).

99. M. Bibikova, et al., High density DNA methylation array with single CpG site resolution. Genomics 98, 288–295 (2011).

100. W. J. Kent, et al., The Human Genome Browser at UCSC. Genome Res 12, 996–1006 (2002).

101. C. Yao, et al., Epigenome-wide association study of whole blood gene expression in Framingham Heart Study participants provides molecular insight into the potential role of CHRNA5 in cigarette smoking-related lung diseases. Clin Epigenetics 13, 1–14 (2021).

102. A. Keshawarz, et al., Expression quantitative trait methylation analysis elucidates gene regulatory effects of DNA methylation: the Framingham Heart Study. Scientific Reports 2023 13:1 13, 1–11 (2023).

103. B. Phipson, J. Maksimovic, A. Oshlack, missMethyl: an R package for analyzing data from Illumina’s HumanMethylation450 platform. Bioinformatics 32, 286–288 (2016).

104. G. Yu, L. G. Wang, Y. Han, Q. Y. He, ClusterProfiler: An R package for comparing biological themes among gene clusters. OMICS 16, 284–287 (2012).

105. T. Battram, et al., The EWAS Catalog: a database of epigenome-wide association studies. Wellcome Open Res 7, 41 (2022).

