## Supplemental Appendix for "Epigenetic Biomarkers of Socioeconomic Status are Associated with Age-Related Chronic Diseases and Mortality in Older Adults"

**This PDF file includes:**

Supplementary text
SI References
Figures S1 to S5
Tables S1 to S19

#### **Section 1**:
#### **Details on Adult SES index components and construction in HRS and MESA**

#### Details on Adult SES index construction

We constructed the adult socioeconomic status (aSES) index by taking the (unweighted) average across all six SES indicators. Detailed information on the six SES measures used to construct the aSES index in HRS and MESA are provided below. For each indicator, we ranked outcomes on a scale from 1 to 5 so that higher values equal more disadvantage as follows: a) Education: 1=>16 years, 2=16 years, 3=13-15 years, 4=12 years, and 5=<12 years; b) Household income: 1=highest income quintile, 5=lowest income quintile; c) Household wealth: 1=highest wealth quintile, 5=lowest wealth quintile; d) Occupational disadvantage: 1=lowest quintile of occupational prestige score, 5=highest quintile of occupational prestige score; e) Neighborhood socioeconomic disadvantage: 1=lowest quintile of score, 5=highest quintile of score; f) Neighborhood social disadvantage: 1=lowest quintile of score, 5=highest quintile of score. We then took the sum of all six elements and divided by the number of non-missing elements (all respondents had at least three elements). Higher index values reflect more disadvantages.

**SES index components**:

- **Education:** In the HRS, education years were coded according to approximate quintiles as follows: 1=>16 years, 2=16 years, 3=13-15 years, 4=12 years, and 5=<12 years. In MESA, information on years of education was not available so we coded the education variable based on highest degree obtained as follows: 1=graduate or professional degree, 2=college degree, 3=some college, 4=high school degree, and 5=no degree.

- **Household income:** In the HRS, we took the average of self-reported, continuous, household income data across all available HRS waves up to 2016 and reverse coded the quintiles as follows: 1=≥$117,465, 2=$74,298-117,464, 3=$48,329-74,297, 4=$27,053-48,328, and 5=$0-27,052. Values were inflation adjusted to 2016 dollars before taking the average across all waves. In MESA, respondents selected their household's income bracket from a series of unfolding brackets rather than reporting a continuous value. We collapsed all 15 income brackets from Exam 5 into approximate household income quintiles for the MESA sample as follows: 1=≥$75,000, 2=$50,000-74,999, 3=$35,000-49,999, 4=$20,000-34,999, and 5=$0-19,999. In both studies, gross household income includes income from respondent and spouse earnings, pensions and annuities, Social Security retirement, unemployment and workers compensation, other government transfers, and household capital income.

- **Household wealth:** In the HRS, we took the average of self-reported, continuous, household wealth data across all available HRS waves up to 2016 and reverse coded the quintiles as follows: 1= ≥$649,583, 2=$286,248-649,582, 3=$118,930-286,247, 4=$27,732-118,929, and 5=$-663,709-27,731. Values were inflation adjusted to 2016 dollars before taking the average across all waves. In MESA, data on household wealth are available in Exam 3. Respondents were asked whether they have any assets in a home, in land, in a car, or in financial investments. Financial investments included assets in stocks, bonds, mutual funds, or retirement investments. We used this information to create five mutually exclusive, dichotomous wealth categories: 1=respondent has assets in a home/land, in a car, and in financial investments; 2=respondent has assets in home/land and one other asset (cars or financial investments); 3=respondent has assets in home/land only or has two other assets (car and financial investments); 4=respondent has assets in a car or in financial investments but does not have assets in home/land; and 5=respondent has no assets.

- **Occupational disadvantage:** We merged occupational prestige scores from the General Social Survey (GSS) with three-digit 1980 or 2000 Census occupation codes in both MESA and the HRS to capture the degree of status associated with a given occupation. To construct a measure of occupational disadvantage, scores were standardized to have a mean of 0 and a standard deviation of 1 and then reverse coded so that higher values reflect less prestige. We used the average occupational disadvantage score across all available exams/waves that MESA/HRS respondents reported working. Occupation data were available for MESA exams 1–3 and across all waves in the HRS up to the 2016 wave.

Occupational prestige scores are weighted average ranks of occupations by samples of workers that have been found to be consistent across raters from different social positions and contexts over time (1-4) and have been found to capture aspects of occupational status related to health and health behaviors that are independent of education and income (5, 6). We used data on occupational prestige from the 2012 GSS (1). A sample of ~1,000 individuals rated 90 occupations each; a rotation of occupation ratings across respondents resulted in ratings for 860 occupational titles. Respondents were asked to rate the social standing of occupations on a hypothetical ladder from top to bottom. Scores adjusted for reviewer-specific effects so that they reflect occupational differences rather than differences between raters using hierarchical linear modelling (HLM).

- **Neighborhood socioeconomic disadvantage score:** We follow past work in MESA that deployed neighborhood-based scores for socioeconomic disadvantage and the social environment created at the Census tract level (where the Census tract is used as a proxy for neighborhoods) (7). The neighborhood socioeconomic disadvantage score for each neighborhood was created based on a factor analysis of 16 census tract level variables from the 2000 Census that reflect dimensions of education, occupation, income, wealth, poverty, employment, and housing. Specifically, the neighborhood socioeconomic disadvantage score is the weighted sum of the following seven standardized variables, which accounted for 49% of the variance in MESA (52% in the HRS) and loaded onto the first factor: 1) percent in census tract with a bachelor's degree; 2) percent with a managerial/professional occupation; 3) percent with a high school education; 4) median home value; 5) median household income; 6) percent with household income greater than 50,000 USD per year; and 7) percent of households with interest, dividends, or net rental income.

  We used the same methodology to create the neighborhood socioeconomic disadvantage score in the HRS using HRS-linked Census data from the HRS-CDR database for items 1)-6) across corresponding Census years (2012-2016) (data on 7 was not available in the HRS-CDR database) (8). Since several years of Census data were used in the HRS we used HRS-based factor weights to construct the score (i.e., factor weights based on the 2000 Census that were used to construct the score in MESA were not applied to the HRS data). Scores were standardized to have a mean of 0 and a standard deviation of 1 for all analysis and reverse coded so that higher values on the scale indicate greater neighborhood socioeconomic disadvantage.

- **Neighborhood social disadvantage score:** We follow past work in MESA that developed a neighborhood social disadvantage score using the sum of conditional empirical Bayes estimate (CEB) scales for aesthetic quality, safety, and social cohesion. The CEB estimates are more reliable than the census-tract crude means because they borrow information from other census tracts in cases where the sample size per tract is very small. Detailed information on the development and validation of this score in MESA is provided elsewhere (9). Briefly, respondents were asked to report their levels of agreement on a 5-point scale (1=strongly agree to 5=strongly disagree) to statements pertaining to neighborhood aesthetic quality, safety, and social cohesion. The CEB estimate for each dimension of the neighborhood social disadvantage score (aesthetic quality, safety, and social cohesion) was estimated from an HLM to account for the fact that each series of questions used to estimate a particular dimension is nested within individuals who are nested within census tracts. The HLM estimates also adjust for site (MESA) or state (HRS) fixed effects, participant age, and sex. The CEB estimates for each dimension were standardized and then aggregated to create the neighborhood social disadvantage score at each MESA/HRS exam/wave.

In MESA, the statements for aesthetic quality were as follows: 1) there is a lot of trash and litter on the street in my neighborhood; 2) there is a lot of noise in my neighborhood; 3) my neighborhood is attractive. The statements for safety were as follows: 1) I feel safe walking in my neighborhood day or night; and 2) violence is a problem in my neighborhood. The statements for social cohesion were as follows: 1) people around here are willing to help their neighbors; 2) people in my neighborhood generally get along with each other; 3) people in my neighborhood can be trusted; and 4) people in my neighborhood share the same values. Where necessary, statements were reverse coded so that higher values indicate worse neighborhood social environment (i.e., lower aesthetic quality, less safety, or lower social cohesion).

In the HRS, we used participant evaluations of their neighborhood environment in the 2006–2016 Psychosocial and Lifestyle Questionnaire (PLQ) that probe how respondents feel about their local area (i.e., everywhere within a 20-minute walk or within a mile of their home) (10). Participants were asked to mark boxes numbered from 1 to 7 on a line, where seven indicated they agree more strongly with the statement. In the HRS PLQ, statements for aesthetic quality were as follows: 1) vandalism and graffiti are a big problem in this area; 2) this area is always full of rubbish and litter; and 3) there are many vacant or deserted houses or storefronts in this area. The statement for safety was as follows (only one measure was available): 1) people would be afraid to walk alone in the area after dark. The statements for social cohesion were as follows: 1) most people in this area cannot be trusted; 2) most people in this area are unfriendly; 3) if you were in trouble, there is nobody in this area who would help you; and 4) I feel that I do not belong to this area.

We used responses from the entire HRS sample to construct Census-tract level estimates of neighborhood socioeconomic and social disadvantages for participants with DNAm data. For both neighborhood scores, we used the cumulative average across all exams/waves to capture average exposure. Scores were standardized to have a mean of 0 and a standard deviation of 1 and reverse coded so that higher values on the scale indicate greater neighborhood social disadvantage.

#### SI References


1. [dataset] Hout M, Smith TW, Marsden PV. *Prestige and Socioeconomic Scores for the 2010 Census Codes*. (2016).
2. Nam CB, Boyd M. Occupational status in 2000; over a century of census-based measurement. *Population Res Policy Rev*. 2004;23:327–358.
3. Smith TW, Son J. Measuring Occupational Prestige on the 2012 General Social Survey. NORC at the University of Chicago. 2014.
4. Treiman DJ. A standard occupational prestige scale for use with historical data. *J Interdisciplinary Hist*. 1976;7:283–304.
5. Barbeau EM, Krieger N, Soobader M-J. Working class matters: socioeconomic disadvantage, race/ ethnicity, gender, and smoking in NHIS 2000. *Am J Public Health*. 2004;94:269–278.
6. Fujishiro K, Roux AVD, Landsbergis P, *et al*. Associations of occupation, job control and job demands with intima-media thickness: the Multi-Ethnic Study of Atherosclerosis (MESA). *Occup Environ Med*. 2011;68:319–326.
7. Roux AVD, Merkin SS, Arnett D, *et al*. Neighborhood of residence and incidence of coronary heart disease. *N Engl J Med*. 2001;345:99–106.
8. [dataset] Ailshire J, Mawhorter S, Choi EY. Contextual Data Resource (CDR): United States Decennial Census and American Community Survey Data, 1990–2018, Version 2.0. Los Angeles, CA: USC/UCLA Center on Biodemography and Population Health. (2020).
9. Mujahid MS, Diez Roux AV, Morenoff JD, et al. Assessing the measurement properties of neighborhood scales: from psychometrics to ecometrics. *Am J Epidemiol*. 2007;165:858–867.
10. Smith J, Ryan L, Fisher GG, et al. Psychosocial and Lifestyle Questionnaire 2006-2016 Documentation Report. Ann Arbor, MI: Survey Research Center, Institute for Social Research. 2017.

### <u>Section 2</u>:
### Supplementary Tables and Figures

**Table S1.** Sample statistics for HRS and MESA

| | HRS | | MESA |
|---|---|---|---|
| | N (%) or Mean (SD) | | |
| | 2016 sample (N=3,527) | 2018 sample (N=3,120) | Exam 5 sample (N=1,182) |
| ***Covariates*** | | | |
| Age | 69.84 (9.68) | 69.58 (9.40) | 69.59 (9.30) |
| Female | 2070 (58.69) | 1855 (59.46) | 601 (50.85) |
| Race | | | |
| White | 2393 (67.85) | 2118 (67.88) | 573 (48.48) |
| Black | 550 (15.59) | 483 (15.48) | 229 (19.37) |
| Hispanic | 481 (13.64) | 428 (13.72) | 380 (32.15) |
| Other | 103 (2.92) | 91 (2.92) | |
| Smoking status | | | |
| Never | 1586 (44.97) | 1436 (46.03) | 574 (48.56) |
| Former | 1563 (44.32) | 1354 (43.40) | 487 (41.20) |
| Current | 378 (10.72) | 330 (10.58) | 121 (10.24) |
| ***Adult SES*** | | | |
| **Adult SES index (range=1-5)** | **3.10 (0.71)** | **3.08 (0.71)** | **2.82 (0.96)** |
| Education years | 12.85 (3.20) | 12.94 (3.20) | |
| Highest degree obtained | | | |
| No degree | 651 (18.51) | 541 (17.40) | 169 (14.32) |
| High school degree | 1094 (31.11) | 954 (30.68) | 228 (19.32) |
| Some college | 870 (24.74) | 778 (25.02) | 388 (32.88) |
| College degree | 476 (13.53) | 442 (14.21) | 186 (15.76) |
| Graduate or professional degree | 426 (12.11) | 395 (12.70) | 209 (17.71) |
| Household Income, thousands of dollars | 82.71 (110.58) | 85.14 (111.79) | |
| Household Income, quintiles (HRS) or given brackets (MESA) | | | |
| $0 - 27,052 (HRS), $0-19,999 (MESA) | 678 (19.22) | 576 (18.46) | 206 (17.93) |
| $27,053 - 48,328 (HRS), $20,000 - 34,999 (MESA) | 714 (20.24) | 602 (19.29) | 236 (20.54) |
| $48,329 - 74,297 (HRS), $35,000 - 49,999 (MESA) | 703 (19.93) | 622 (19.94) | 222 (19.32) |
| $74,298 - 117,464 (HRS), $50,000 - 74,999 (MESA) | 731 (20.73) | 661 (21.19) | 215 (18.71) |
| $117,465+ (HRS), $75,000+ (MESA) | 701 (19.88) | 659 (21.12) | 270 (23.50) |
| Occupational disadvantage | -0.05 (0.97) | -0.06 (0.97) | -0.00 (1.00) |
| Neighborhood socioeconomic disadvantage | -0.43 (4.79) | -0.35 (4.85) | 0.02 (4.87) |
| Neighborhood social environment | -0.14 (2.34) | -0.16 (2.37) | 0.42 (2.68) |
| Wealth | | | |
| Household wealth, thousands of dollars | 455.22 (928.58) | 466.46 (934.76) | |
| Household wealth, quintiles (HRS) or assets (MESA) | | | |
| $-663,709 - 27,731 | 671 (19.03) | 592 (18.97) | |
| $27,732 - 118,929 | 710 (20.14) | 605 (19.39) | |
| $118,930 - 286,247 | 703 (19.94) | 617 (19.78) | |
| $286,248 - 649,582 | 730 (20.70) | 653 (20.93) | |
| $649,583+ | 712 (20.19) | 653 (20.93) | |
| No assets | | | 112 (9.60) |
| Assets in financial investments or car and not home/land | | | 134 (11.48) |
| Assets in home/land only or car and investments | | | 83 (7.11) |
| Assets in home/land and car or investments | | | 178 (15.25) |
| Assets in home/land, car, and financial investments | | | 660 (56.56) |
| ***Childhood SES*** | | | |
| **Childhood SES index (range=0-4)** | **1.70 (0.89)** | **1.68 (0.89)** | **2.35 (0.99)** |
| Parental education, years | 10.87 (3.92) | 10.95 (3.92) | |
| Mother’s education, years | 9.97 (3.98) | 10.04 (3.99) | |

| | | | |
|---|---|---|---|
| Mother's education | | | |
| <8 years (HRS), no schooling (MESA) | 786 (22.29) | 676 (21.67) | 87 (7.55) |
| 8 to 11 years (HRS), some school without HS degree (MESA) | 958 (27.17) | 834 (26.74) | 530 (45.97) |
| 12 years (HRS), HS degree (MESA) | 1172 (33.24) | 1047 (33.57) | 324 (28.10) |
| 13 to 15 years (HRS), some college without degree (MESA) | 326 (9.25) | 297 (9.52) | 92 (7.98) |
| 16+ years (HRS), college degree/graduate school (MESA) | 284 (8.05) | 265 (8.50) | 120 (10.41) |
| Father's education, years | 9.68 (4.22) | 9.75 (4.23) | |
| Father's education | | | |
| <8 years (HRS), no schooling (MESA) | 939 (26.64) | 814 (26.11) | 94 (8.32) |
| 8 to 11 years (HRS), some school without HS degree (MESA) | 1020 (28.94) | 883 (28.32) | 563 (49.82) |
| 12 years (HRS), HS degree (MESA) | 900 (25.53) | 811 (26.01) | 256 (22.65) |
| 13 to 15 years (HRS), some college without degree (MESA) | 302 (8.57) | 270 (8.66) | 90 (7.96) |
| 16+ years (HRS), college degree/graduate school (MESA) | 364 (10.33) | 340 (10.90) | 127 (11.24) |
| Childhood financial strain index (range=0-4) | 0.92 (1.16) | 0.91 (1.16) | |
| ***Epigenetic aging measures*** | | | |
| GrimAge | 68.37 (8.65) | 67.95 (8.38) | 79.37 (8.07) |
| Dunedin Pace of Aging (DunedinPACE) | 1.04 (0.15) | 1.03 (0.14) | 1.29 (0.11) |
| ***Health conditions*** | | | |
| Number of chronic conditions | | 2.58 (1.54) | |
| Cardiometabolic conditions index | | 0.01 (1.00) | 0.00 (1.00) |
| Self-reported health | | 2.89 (1.04) | 2.59 (0.90) |
| Mortality | | 162 (4.59) | 184 (15.59) |
| Langa-Weir dementia (HRS), ICD all cause dementia (MESA) | | 136 (4.36) | 67 (5.68) |

**Notes:** The childhood SES index could not be constructed in MESA because childhood financial information was not available. Information on health conditions in MESA was taken from Exam 6. HRS values for household income and wealth are in 2016 dollars. MESA values for household income are in 2012 dollars.

**Figure S1.** Manhattan plot of adult SES index EWAS results in HRS

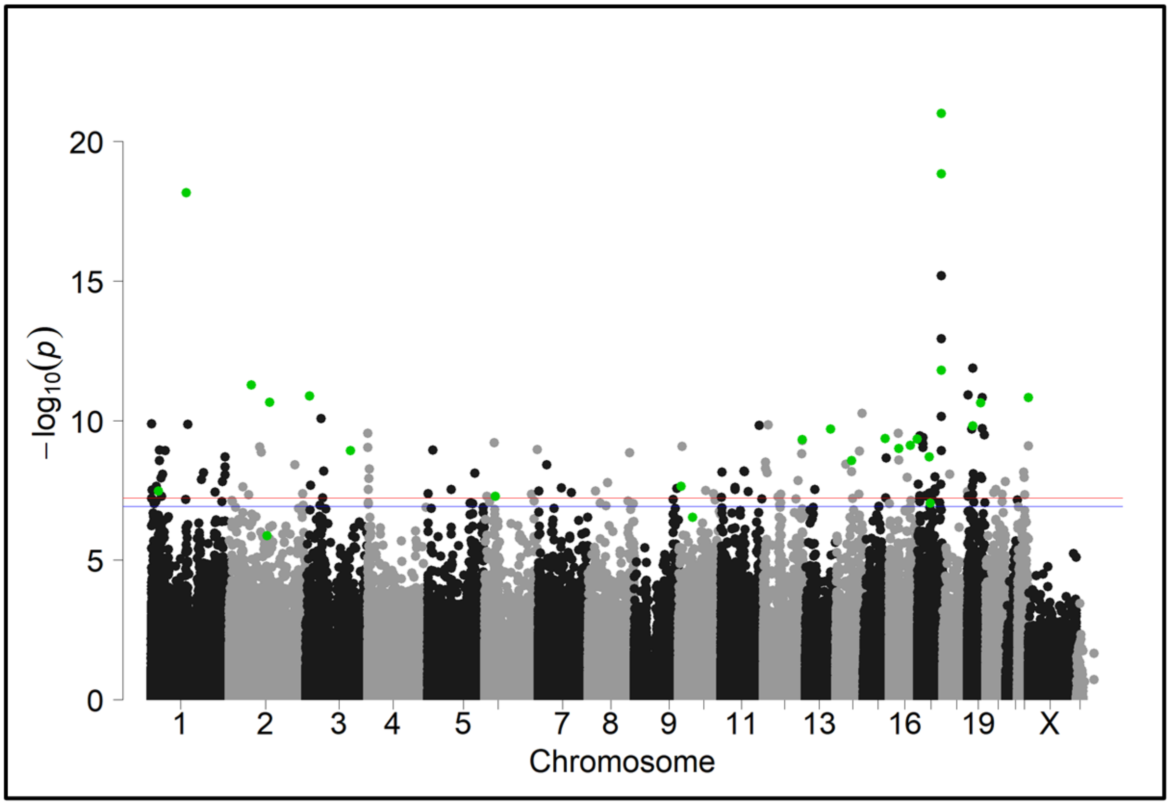


**Notes:** EWAS Model: CpG = adult SES Index + age + sex + race + smoking + WBCs + genetic PC1-PC4 + technical covariates (random effect).

1.2e-7 ~ FDR=0.1

6e-8 ~ FDR=0.05

Enet selected sites

**Figure S2.** Manhattan plot of childhood SES index EWAS results in HRS

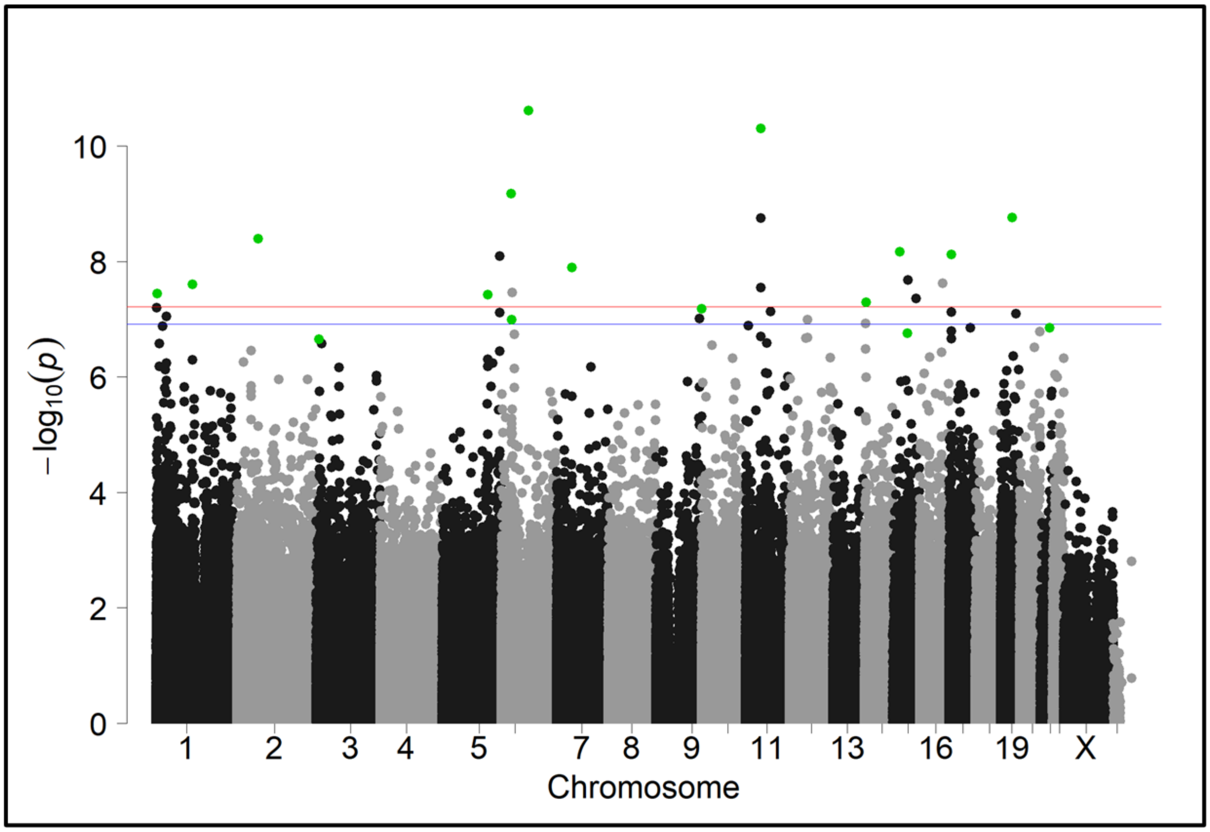


**Notes:** EWAS Model: CpG = childhood SES Index + age + sex + race + smoking + WBCs + genetic PC1-PC4 + technical covariates (random effect).

1.2e-7 ~ FDR=0.1

6e-8 ~ FDR=0.05

Enet selected sites

**Table S2.** EWAS p-values of adult SES index components in HRS

| CpG | P aSES | aSES P rank | P educ | P hhinc | P wealth | P neighdis | P socenv | P occ_prest |
|---|---|---|---|---|---|---|---|---|
| cg18181703 | 9.81E-22 | 1 | **2.68E-10** | **6.36E-13** | **4.43E-15** | **9.95E-11** | 2.33E-05 | **1.02E-06** |
| cg11047325 | 1.43E-19 | 2 | **2.79E-09** | **7.22E-13** | **4.20E-15** | **1.47E-10** | 1.58E-05 | 2.69E-03 |
| cg21327712 | 6.90E-19 | 3 | **9.56E-12** | **2.25E-11** | **1.88E-12** | **7.06E-10** | 1.66E-02 | **1.64E-07** |
| cg01894508 | 5.30E-12 | 7 | 2.24E-05 | **1.25E-10** | **4.98E-08** | **6.94E-07** | 9.45E-03 | 1.05E-03 |
| cg18062721 | 1.34E-11 | 9 | **1.39E-07** | 8.42E-04 | **5.53E-09** | **1.73E-07** | 8.85E-04 | 2.65E-04 |
| cg03362418 | 1.51E-11 | 11 | **7.72E-08** | **2.36E-09** | **1.02E-09** | 1.63E-04 | 3.16E-02 | 4.09E-03 |
| cg25008217 | 2.28E-11 | 12 | **2.38E-07** | 3.51E-05 | 1.02E-04 | **2.49E-07** | 6.14E-03 | **3.57E-07** |
| cg17461390 | 1.58E-10 | 19 | **1.38E-06** | 1.48E-05 | **2.49E-07** | **6.47E-08** | 2.70E-03 | 6.10E-02 |
| cg02767093 | 2.00E-10 | 21 | 6.38E-05 | 2.14E-05 | **6.56E-08** | 2.00E-05 | 5.33E-03 | 1.05E-04 |
| cg25145728 | 4.54E-10 | 28 | **3.80E-07** | 2.76E-05 | 1.95E-05 | 1.09E-05 | 3.78E-03 | 8.40E-02 |
| cg02017926 | 4.93E-10 | 29 | 1.41E-05 | 5.29E-05 | 2.44E-05 | **3.27E-09** | 2.14E-02 | 2.50E-03 |
| cg10922280 | 7.85E-10 | 33 | 1.46E-04 | **3.31E-06** | **5.64E-07** | 2.03E-05 | 5.56E-03 | 4.42E-05 |
| cg10206344 | 9.93E-10 | 39 | 1.11E-05 | **6.22E-06** | **2.45E-06** | **8.06E-06** | 4.75E-03 | 6.74E-03 |
| cg11454468 | 1.21E-09 | 44 | **4.48E-06** | 6.59E-03 | 4.19E-05 | **1.69E-06** | 7.18E-05 | 3.47E-03 |
| cg11183072 | 2.04E-09 | 48 | **1.28E-09** | 3.94E-04 | 6.80E-04 | 5.83E-05 | 3.35E-02 | 3.05E-04 |
| cg10842530 | 2.70E-09 | 52 | 1.39E-04 | **2.09E-06** | 2.06E-04 | 1.47E-05 | 3.69E-04 | 2.08E-04 |
| cg03031609 | 2.31E-08 | 82 | **3.15E-06** | **5.90E-06** | 1.44E-04 | 1.31E-05 | 1.18E-01 | 1.36E-03 |
| cg24445316 | 3.43E-08 | 95 | 1.29E-05 | 1.13E-05 | 3.29E-03 | 1.66E-04 | 1.17E-02 | 2.01E-04 |
| cg06864083 | 9.04E-08 | 142 | 5.60E-04 | 9.06E-05 | 2.61E-04 | 2.51E-04 | 8.60E-02 | **8.98E-06** |
| cg03090734 | 2.98E-07 | 205 | 5.31E-05 | **9.86E-06** | 1.47E-05 | 6.47E-03 | 1.13E-01 | 1.63E-03 |
| cg10946295 | 1.34E-06 | 373 | 1.34E-03 | 1.23E-04 | 7.49E-04 | 1.64E-02 | 2.00E-03 | 5.79E-03 |
| cg27637521 | 4.22E-06 | 576 | 2.38E-05 | **2.17E-12** | **3.57E-09** | **9.98E-10** | 3.40E-02 | 2.13E-03 |
| cg06021088 | 6.67E-06 | 687 | **1.70E-06** | **3.06E-07** | **1.75E-09** | **1.38E-06** | 3.53E-03 | 2.33E-02 |
| cg19574915 | 1.45E-04 | 2698 | 9.13E-04 | 3.79E-05 | **8.64E-06** | **2.07E-09** | 2.55E-04 | 2.90E-04 |
| cg08852765 | 4.57E-03 | 15540 | 1.51E-05 | 1.06E-03 | **5.31E-06** | **1.98E-06** | 5.91E-02 | 1.71E-02 |

**Notes**: CpG = Elastic Net selected CpG sites for adult SES index biomarker; P aSES = EWAS p-value for adult SES; aSES P rank = Ordered p-value rank of CpG site in adult SES EWAS; P educ = EWAS p-value for education years (quintiles); P hhinc = EWAS p-value for household income (quintiles); P wealth = EWAS p-value for total wealth assets (quintiles); P neighdis = EWAS p-value for neighborhood disadvantage score (quintiles); P socenv = EWAS p-value for social environment score (quintiles); P occ_prest = EWAS p-value for occupational disadvantage score (quintiles). P-values < 1.0E-5 are bolded.

**Table S3.** EWAS p-values of childhood SES index components in HRS

| CpG | P cSES | cSES P rank | P peduc | P cfsi |
|---|---|---|---|---|
| cg03519157 | 1.96E-11 | 1 | **2.04E-10** | 7.43E-05 |
| cg04887278 | 3.20E-11 | 2 | **3.09E-09** | 4.48E-05 |
| cg08469255 | 6.55E-10 | 3 | **2.00E-06** | 1.37E-05 |
| cg22505924 | 1.91E-09 | 5 | **4.94E-09** | 1.31E-03 |
| cg16329896 | 2.96E-09 | 6 | **1.33E-07** | 1.27E-03 |
| cg27496526 | 3.00E-09 | 7 | **8.48E-07** | 9.71E-05 |
| cg17810176 | 4.70E-09 | 8 | **8.99E-08** | 4.05E-04 |
| cg25563256 | 1.02E-08 | 9 | **1.55E-10** | 8.56E-03 |
| cg12452298 | 1.15E-08 | 11 | 1.12E-05 | 1.68E-04 |
| cg10394832 | 2.33E-08 | 14 | **2.83E-06** | 9.46E-05 |
| cg21159993 | 2.51E-08 | 15 | **1.17E-08** | 7.53E-03 |
| cg17496659 | 3.30E-08 | 17 | **2.01E-10** | 1.29E-02 |
| cg02170695 | 1.67E-07 | 36 | 9.19E-02 | **2.65E-09** |
| cg05302489 | 1.79E-07 | 38 | 2.38E-02 | **5.58E-08** |
| cg13067434 | 3.28E-07 | 52 | 1.83E-04 | 3.70E-05 |

**Notes:** CpG = Elastic Net selected CpG sites for childhood SES index biomarker; P cSES = EWAS p-value for childhood SES; cSES P rank is the ordered p-value rank of the CpG site in the childhood SES EWAS; P peduc = EWAS p-value for parental education (quintiles); P cfsi = EWAS p-value for childhood financial strain index (quintiles). P-values < 1.0E-5 are bolded.

**Table S4.** CpG sites selected for 450K derived adult SES index biomarker (aSES-BIO450) by elastic net in HRS sample

| CpG | E-Net Weight | Chr. | Pos. | N | EWAS beta | EWAS p-value | EWAS rank | UCSC RefGene Name | UCSC RefGene Group | Relation to UCSC CpG Island | Regulatory Feature Group | Phantom5 Enhancer | DHS | eQTM |
|---|---|---|---|---|---|---|---|---|---|---|---|---|---|---|
| cg18181703 | -0.9083 | 17 | 76354621 | 3522 | -0.011 | 9.80E-22 | 1 | *SOCS3* | Body | N_Shore | Promoter associated | 0 | 0 | 1 |
| cg01894508 | 0.2937 | 2 | 70189111 | 3517 | 0.007 | 5.30E-12 | 7 | *ASPRV1* | 5'UTR; 1stExon | S_Shore | | 0 | 0 | 0 |
| cg18062721 | 0.1507 | 3 | 11643427 | 3518 | 0.013 | 1.30E-11 | 9 | *VGLL4* | Body | | | 0 | 1 | 0 |
| cg01406381 | -0.1363 | 19 | 47288263 | 3520 | -0.003 | 1.50E-11 | 10 | *SLC1A5* | TSS200; Body; 5'UTR | N_Shelf | Promoter associated | 0 | 1 | 1 |
| cg25691553 | -0.223 | 3 | 49057884 | 3526 | -0.002 | 8.40E-11 | 15 | *NDUFAF3; DALRD3; MIR425* | TSS200; 5'UTR; TSS1500 | N_Shore | Promoter associated | 0 | 1 | 1 |
| cg02767093 | -0.2555 | 13 | 99130655 | 3480 | -0.004 | 2.00E-10 | 21 | *STK24* | Body | S_Shelf | | 1 | 1 | 1 |
| cg24704287 | -0.0004 | 19 | 13951481 | 3517 | -0.007 | 2.10E-10 | 22 | | | N_Shore | Promoter associated | 0 | 1 | 1 |
| cg19769182 | 0.0344 | 16 | 29823868 | 3520 | 0.005 | 2.90E-10 | 23 | *PRRT2* | 5'UTR | Island | | 0 | 1 | 1 |
| cg02017926 | 0.0464 | 12 | 1.24E+08 | 3517 | 0.006 | 4.90E-10 | 29 | *CDK2AP1* | Body | Island | Promoter associated | 0 | 1 | 1 |
| cg08696931 | 0.0157 | 12 | 1.24E+08 | 3512 | 0.005 | 5.20E-10 | 30 | *CDK2AP1* | Body | Island | | 0 | 1 | 1 |
| cg08469255 | 0.0167 | 6 | 30851069 | 3451 | 0.005 | 6.30E-10 | 31 | *DDR1* | TSS1500 | N_Shore | Unclassified | 0 | 1 | 1 |
| cg10922280 | 0.1476 | 16 | 68034227 | 3517 | 0.005 | 7.90E-10 | 33 | *DPEP2* | TSS1500 | | | 0 | 1 | 1 |
| cg10206344 | 0.2541 | 16 | 31483277 | 3525 | 0.003 | 9.90E-10 | 39 | *TGFB1I1* | TSS1500; TSS200 | Island | Unclassified | 0 | 1 | 0 |
| cg25130381 | 0.0093 | 1 | 27440721 | 3451 | 0.005 | 1.10E-09 | 41 | *SLC9A1* | Body | | | 0 | 1 | 1 |
| cg11183072 | 0.085 | 17 | 37894397 | 3522 | 0.005 | 2.00E-09 | 48 | *GRB7* | TSS200; 5'UTR | | | 0 | 1 | 0 |
| cg01101459 | 0.0245 | 1 | 2.35E+08 | 3513 | 0.007 | 4.60E-09 | 56 | | | | | 0 | 0 | 0 |
| cg08136809 | 0.1109 | 19 | 41882642 | 3524 | 0.008 | 1.00E-08 | 68 | *TMEM91* | 1stExon; 5'UTR; TSS1500 | Island | | 0 | 1 | 0 |
| cg11846112 | 0.0003 | 1 | 2.28E+08 | 3509 | 0.006 | 1.60E-08 | 76 | | | Island | Unclassified | 0 | 1 | 1 |
| cg03031609 | 0.0932 | 10 | 7453871 | 3521 | 0.003 | 2.30E-08 | 82 | *SFMBT2* | TSS1500 | Island | | 0 | 1 | 1 |
| cg18385254 | 0.0666 | 18 | 77724395 | 3517 | 0.004 | 3.60E-08 | 97 | *HSBP1L1* | TSS200 | Island | Promoter associated | 0 | 0 | 0 |

| | | | | | | | | | | | | | | |
|---|---|---|---|---|---|---|---|---|---|---|---|---|---|---|
| cg27491190 | 0.036 | 12 | 46767943 | 3522 | 0.005 | 5.50E-08 | 116 | *SLC38A2* | TSS1500 | S_Shore | Promoter associated | 0 | 1 | 1 |
| cg06864083 | 0.1675 | 17 | 40832319 | 3520 | 0.004 | 9.00E-08 | 142 | *CCR10* | Body | Island | | 0 | 1 | 0 |
| cg26504305 | 0.0181 | 19 | 19281019 | 3517 | 0.007 | 3.90E-07 | 232 | *LOC729991-MEF2B; MEF2B* | Body; 5'UTR; 1stExon | Island | Unclassified | 0 | 0 | 0 |
| cg27637521 | -0.412 | 17 | 76355202 | 3519 | -0.002 | 4.20E-06 | 576 | *SOCS3* | 5'UTR | Island | | 0 | 1 | 1 |
| cg22488164 | 0.0422 | 12 | 14716910 | 3524 | 0.007 | 4.70E-06 | 598 | *PLBD1* | Body | N_Shelf | | 0 | 0 | 1 |
| cg15866367 | 0.0114 | 13 | 49796387 | 3518 | 0.005 | 6.60E-06 | 681 | *MLNR* | Body | S_Shore | | 0 | 1 | 0 |
| cg06021088 | -0.1431 | 2 | 1.28E+08 | 3515 | -0.004 | 6.70E-06 | 687 | *BIN1* | Body | | Unclassified | 0 | 1 | 1 |
| cg19574915 | 0.1898 | 15 | 89195555 | 3526 | 0.004 | 1.50E-04 | 2698 | *ISG20* | Body | | Promoter associated cell type specific | 0 | 1 | 1 |
| cg08852765 | 0.0889 | 6 | 33396407 | 3513 | 0.002 | 4.60E-03 | 15540 | *SYNGAP1* | Body | S_Shore | Promoter associated | 1 | 1 | 0 |

**Notes:** Chr., chromosome; EWAS, epigenome-wide association study; N_Shore, north shore; Pos., position; S_Shore, south shore, UCSC, Univesity of California Santa Cruz Genome Browser; DHS, DNAse hypersensitive site; eQTM, expression quantitative trait methylation site.

**Table S5.** CpG sites selected for 450K derived childhood SES index biomarker (cSES-BIO450) by elastic net in HRS sample

| CpG | E-Net Weight | Chr. | Pos. | N | EWAS beta | EWAS p-value | EWAS rank | UCSC RefGene Name | UCSC RefGene Group | Relation to UCSC CpG Island | Regulatory Feature Group | Phantom5 Enhancer | DHS | eQTM |
|---|---|---|---|---|---|---|---|---|---|---|---|---|---|---|
| cg04887278 | 0.5964 | 6 | 83903927 | 3505 | 0.004 | 2.40E-11 | 1 | *PGM3; RWDD2A* | TSS1500; 5'UTR | S_Shore | | 0 | 1 | 1 |
| cg03519157 | 0.4842 | 11 | 46367033 | 3526 | 0.004 | 4.90E-11 | 2 | *DGKZ* | 5'UTR; 1stExon; Body | Island | | 0 | 1 | 1 |
| cg08469255 | 0.2306 | 6 | 30851069 | 3451 | 0.006 | 6.70E-10 | 3 | *DDR1* | TSS1500 | N_Shore | Unclassified | 0 | 1 | 1 |
| cg17810176 | 0.1133 | 19 | 36035831 | 3523 | 0.006 | 1.70E-09 | 4 | *GAPDHS; TMEM147* | Body; TSS1500 | N_Shore | | 0 | 1 | 1 |
| cg27496526 | 0.4926 | 15 | 41805530 | 3526 | 0.002 | 6.90E-09 | 7 | *LTK* | Body | Island | | 0 | 1 | 0 |
| cg25563256 | 0.0424 | 17 | 7341641 | 3524 | 0.006 | 7.70E-09 | 8 | *FGF11* | TSS1500 | N_Shore | Unclassified | 0 | 1 | 1 |
| cg16329896 | 0.1317 | 7 | 47515060 | 3517 | 0.004 | 1.30E-08 | 10 | *TNS3* | 5'UTR | | Unclassified cell type specific | 1 | 1 | 1 |
| cg12452298 | 0.0011 | 15 | 67134587 | 3524 | 0.005 | 2.10E-08 | 11 | | | N_Shore | Unclassified cell type specific | 0 | 1 | 1 |
| cg10394832 | 0.0682 | 1 | 1.12E+08 | 3525 | 0.007 | 2.50E-08 | 13 | *C1orf88* | Body | Island | Promoter associated | 0 | 0 | 0 |
| cg17496659 | 0.0202 | 1 | 3.57E+06 | 3520 | 0.006 | 3.60E-08 | 16 | *TP73* | TSS1500 | Island | | 0 | 1 | 1 |
| cg21159993 | 0.0298 | 5 | 1.40E+08 | 3523 | 0.004 | 3.70E-08 | 17 | *C5orf32* | TSS1500 | Island | Promoter associated | 0 | 1 | 0 |
| cg19989295 | 0.0054 | 14 | 24641077 | 3483 | 0.004 | 5.10E-08 | 19 | *REC8* | TSS200 | Island | Promoter associated | 0 | 1 | 0 |
| cg13825574 | 0.0334 | 9 | 1.40E+08 | 3514 | 0.003 | 6.60E-08 | 21 | *RNF208* | TSS1500 | Island | | 0 | 1 | 0 |
| cg05302489 | 0.0442 | 6 | 31760426 | 3499 | 0.008 | 1.00E-07 | 28 | *VARS* | Body | N_Shelf | | 0 | 0 | 1 |
| cg12748367 | 0.1059 | 2 | 2.16E+08 | 3524 | 0.001 | 1.10E-06 | 82 | *FN1* | TSS1500 | S_Shore | Unclassified | 0 | 1 | 0 |

**Notes:** Chr., chromosome; EWAS, epigenome-wide association study; N_Shore, north shore; Pos., position; S_Shore, south shore, UCSC, Univesity of California Santa Cruz Genome Browser; DHS, DNAse hypersensitive site; eQTM, expression quantitative trait methylation site.

**Table S6.** Correlations between SES indices, SES biomarkers, and epigenetic aging measures in HRS

| | Adult SES index biomarker (aSES-BIO) | Childhood SES index biomarker (cSES-BIO) | Adult SES index biomarker 450K (aSES-BIO450) | Childhood SES index biomarker 450K (cSES-BIO450) | Adult SES index (aSES) | Childhood SES index (cSES) | GrimAge | Dunedin-PACE |
|---|---|---|---|---|---|---|---|---|
| Adult SES index biomarker (aSES-BIO) | 1.000 | | | | | | | |
| Childhood SES index biomarker (cSES-BIO) | **0.284** | 1.000 | | | | | | |
| Adult SES index biomarker 450K (aSES-BIO450) | **0.888** | **0.261** | 1.000 | | | | | |
| Childhood SES index biomarker 450K (cSES-BIO450) | **0.270** | **0.988** | **0.246** | 1.000 | | | | |
| Adult SES index (aSES) | **0.235** | **0.107** | **0.230** | **0.106** | 1.000 | | | |
| Childhood SES index (cSES) | **0.118** | **0.190** | **0.115** | **0.181** | **0.429** | 1.000 | | |
| GrimAge | **0.180** | **0.049** | **0.195** | **0.044** | **0.055** | **0.158** | 1.000 | |
| DunedinPACE | **0.497** | **0.162** | **0.512** | **0.141** | **0.320** | **0.201** | **0.419** | 1.000 |

**Notes:** Pearson correlation coefficients with p<.05 under H0: r=0 are bolded. N=3527

**Table S7.** Associations between SES biomarkers and health outcomes in the HRS

| | | **Adult SES** | | | | | | | |
|---|---|---|---|---|---|---|---|---|---|
| | | **aSES** | **aSES-BIO** | | | | | | |
| Outcome | Estimate Type | Model 1* | Model 1 | Model 2 | Model 3a | Model 4a | Model 3b | Model 4b | Model 5 |
| Number of chronic conditions | IRR | 1.17*** (1.14, 1.20) | 1.12*** (1.10, 1.15) | 1.09*** (1.07, 1.12) | 1.08*** (1.05, 1.11) | 1.06*** (1.03, 1.08) | 1.05** (1.02, 1.08) | 1.03** (1.00, 1.05) | 1.11*** (1.09, 1.14) |
| Cardiometabolic conditions index | Beta | 0.21*** (0.17, 0.25) | 0.18*** (0.15, 0.21) | 0.14*** (0.11, 0.18) | 0.12*** (0.08, 0.16) | 0.09*** (0.05, 0.13) | 0.06** (0.02, 0.10) | 0.04* (-0.004, 0.07) | 0.17*** (0.14, 0.20) |
| Self-reported health status | Beta | 0.32*** (0.29, 0.36) | 0.18*** (0.15, 0.22) | 0.12*** (0.09, 0.16) | 0.13*** (0.09, 0.16) | 0.07** (0.04, 0.11) | 0.10*** (0.06, 0.14) | 0.05** (0.01, 0.09) | 0.17*** (0.14, 0.21) |
| Mortality | OR | 1.49*** (1.23, 1.80) | 1.62*** (1.38, 1.90) | 1.55*** (1.31, 1.83) | 1.30** (1.09, 1.56) | 1.25** (1.05, 1.50) | 1.33** (1.10, 1.61) | 1.29** (1.06, 1.56) | 1.60*** (1.36, 1.88) |
| Langa-Weir dementia | OR | 1.96*** (1.53, 2.50) | 1.26** (1.05, 1.52) | 1.14 (0.94, 1.38) | 1.22* (1.00, 1.49) | 1.11 (0.91, 1.37) | 1.27** (1.03, 1.58) | 1.18 (0.95, 1.46) | 1.23** (1.02, 1.48) |
| | | **Childhood SES** | | | | | | | |
| | | **cSES** | **cSES-BIO** | | | | | | |
| Outcome | Estimate Type | Model 1* | Model 1 | Model 2 | Model 3a | Model 4a | Model 3b | Model 4b | Model 5 |
| Number of chronic conditions | IRR | 1.10*** (1.07, 1.12) | 1.05*** (1.03, 1.07) | 1.03** (1.01, 1.05) | 1.04** (1.01, 1.06) | 1.02* (1.00, 1.04) | 1.02** (1.00, 1.04) | 1.01 (0.99, 1.03) | 1.03** (1.01, 1.05) |
| Cardiometabolic conditions index | Beta | 0.12*** (0.09, 0.15) | 0.05** (0.02, 0.09) | 0.03** (0.0004, 0.07) | 0.04** (0.00, 0.07) | 0.02 (-0.02, 0.05) | 0.01 (-0.02, 0.05) | -0.003 (-0.04, 0.03) | 0.03** (0.001, 0.07) |
| Self-reported health | Beta | 0.14*** (0.10, 0.17) | 0.07*** (0.04, 0.10) | 0.05** (0.01, 0.08) | 0.05** (0.02, 0.09) | 0.03* (-0.003, 0.07) | 0.04** (0.00, 0.07) | 0.02 (-0.02, 0.05) | 0.04** (0.01, 0.07) |
| Mortality | OR | 1.24** (1.05, 1.46) | 1.25** (1.07, 1.45) | 1.21** (1.04, 1.41) | 1.17** (1.01, 1.37) | 1.14* (0.98, 1.33) | 1.15* (0.99, 1.33) | 1.12 (0.96, 1.30) | 1.21** (1.04, 1.41) |
| Langa-Weir dementia | OR | 1.46** (1.20, 1.77) | 1.10 (0.93, 1.31) | 1.03 (0.86, 1.23) | 1.09 (0.92, 1.30) | 1.02 (0.86, 1.22) | 1.09 (0.92, 1.30) | 1.02 (0.85, 1.22) | 1.06 (0.89, 1.26) |

**Notes:** Table displays estimates from separate regressions of health or mortality on aSES-BIO or cSES-BIO. The aSES-BIO and cSES-BIO measures were standardized for analysis. Model 1 controls for age, sex, race, current smoking, former smoking, and *APOE* e4 carrier status (dementia models only). Model 2 adds controls for measured aSES (in aSES-BIO regressions) or cSES (in cSES-BIO regressions). Models 3a and 4a add GrimAge to Model 1 (3a) or Model 2 (4a). Models 3b and 4b add DunedinPACE to Model 1 (3b) or Model 2 (4b). Model 5 adds measured cSES (for adult SES models) or measured aSES (for childhood SES models). Model 1* is identical to Model 1 but replaces the biomarkers with measured aSES or cSES. For chronic conditions, a Poisson model was used; for the cardiometabolic conditions index (CMCI) and self-reported health status (SRHS) linear models were used; for mortality and dementia logistic models were used. 95% confidence intervals in parentheses. IRR, incidence rate ratio; OR, odds ratio. Sample sizes: number of chronic conditions=3120; CMCI=3120; SRHS=3118; mortality=3527; dementia=3120.

**Table S8.** Associations between 450k SES biomarkers and health outcomes in the HRS

| | | Adult SES | | | | | | | |
|---|---|---|---|---|---|---|---|---|---|
| | | aSES | aSES-BIO450 | | | | | | |
| Outcome | Estimate Type | Model 1* | Model 1 | Model 2 | Model 3a | Model 4a | Model 3b | Model 4b | Model 5 |
| Number of chronic conditions | IRR | 1.17***<br>(1.14, 1.20) | 1.12***<br>(1.10, 1.15) | 1.09***<br>(1.07, 1.12) | 1.08***<br>(1.05, 1.11) | 1.06***<br>(1.03, 1.08) | 1.05**<br>(1.02, 1.08) | 1.03**<br>(1.00, 1.06) | 1.12***<br>(1.09, 1.14) |
| Cardiometabolic conditions index | Beta | 0.21***<br>(0.17, 0.25) | 0.19***<br>(0.15, 0.22) | 0.15***<br>(0.11, 0.18) | 0.12***<br>(0.08, 01.16) | 0.09***<br>(0.05, 0.13) | 0.06**<br>(0.02, 0.10) | 0.04*<br>(-0.003, 0.08) | 0.18***<br>(0.14, 0.21) |
| Self-reported health | Beta | 0.32***<br>(0.29, 0.36) | 0.20***<br>(0.16, 0.23) | 0.14***<br>(0.10, 0.18) | 0.14***<br>(0.10, 0.18) | 0.09***<br>(0.05, 0.13) | 0.12***<br>(0.07, 0.16) | 0.07**<br>(0.03, 0.11) | 0.19***<br>(0.15, 0.22) |
| Mortality | OR | 1.49***<br>(1.23, 1.80) | 1.73***<br>(1.48, 2.03) | 1.67***<br>(1.41, 1.97) | 1.40**<br>(1.17, 1.67) | 1.35**<br>(1.12, 1.62) | 1.46***<br>(1.21, 1.77) | 1.42**<br>(1.17, 1.72) | 1.17***<br>(1.46, 2.01) |
| Langa-Weir Dementia | OR | 1.96***<br>(1.53, 2.50) | 1.27**<br>(1.06, 1.54) | 1.15<br>(0.95, 1.39) | 1.23**<br>(1.01, 1.51) | 1.12<br>(0.91, 1.39) | 1.29**<br>(1.04, 1.61) | 1.19<br>(0.95, 1.49) | 1.24**<br>(1.03, 1.50) |
| | | **Childhood SES** | | | | | | | |
| | | **cSES** | **cSES-BIO450** | | | | | | |
| Outcome | Estimate Type | Model 1* | Model 1 | Model 2 | Model 3a | Model 4a | Model 3b | Model 4b | Model 5 |
| Number of chronic conditions | IRR | 1.10***<br>(1.07, 1.12) | 1.05***<br>(1.02, 1.07) | 1.03**<br>(1.01, 1.05) | 1.04**<br>(1.01, 1.06) | 1.02*<br>(1.00, 1.04) | 1.02**<br>(1.00, 1.05) | 1.01<br>(0.99, 1.03) | 1.03**<br>(1.01, 1.05) |
| Cardiometabolic conditions index | Beta | 0.12***<br>(0.09, 0.15) | 0.05**<br>(0.02, 0.09) | 0.03*<br>(-0.002, 0.07) | 0.03**<br>(0.002, 0.07) | 0.02<br>(-0.02, 0.05) | 0.02<br>(-0.02, 0.05) | 0.0001<br>(-0.03, 0.03) | 0.03*<br>(-0.002, 0.06) |
| Self-reported health | Beta | 0.14***<br>(0.10, 0.17) | 0.07***<br>(0.04, 0.11) | 0.05**<br>(0.02, 0.09) | 0.06**<br>(0.02, 0.09) | 0.04**<br>(0.004, 0.07) | 0.04**<br>(0.01, 0.08) | 0.03<br>(-0.01, 0.06) | 0.04**<br>(0.01, 0.08) |
| Mortality | OR | 1.24**<br>(1.05, 1.46) | 1.26**<br>(1.08, 1.46) | 1.22**<br>(1.05, 1.43) | 1.19**<br>(1.03, 1.39) | 1.16*<br>(0.99, 1.35) | 1.17**<br>(1.01, 1.36) | 1.14*<br>(0.98, 1.33) | 1.22**<br>(1.05, 1.42) |
| Langa-Weir Dementia | OR | 1.46**<br>(1.20, 1.77) | 1.09<br>(0.92, 1.30) | 1.02<br>(0.86, 1.22) | 1.08<br>(0.91, 1.29) | 1.01<br>(0.85, 1.21) | 1.08<br>(0.91, 1.29) | 1.02<br>(0.85, 1.21) | 1.05<br>(0.88, 1.25) |

**Notes:** Table displays estimates from separate regressions of health or mortality on aSES-BIO450 or cSES-BIO450. The aSES-BIO and cSES-BIO measures were standardized for analysis. Model 1 controls for age, sex, race, current smoking, former smoking, and *APOE* e4 carrier status (dementia models only). Model 2 adds controls for measured aSES (in aSES-BIO450 regressions) or cSES (in cSES-BIO450 regressions). Models 3a and 4a add GrimAge to Model 1 (3a) or Model 2 (4a). Models 3b and 4b add DunedinPACE to Model 1 (3b) or Model 2 (4b). Model 5 adds measured cSES (for adult SES models) or measured aSES (for childhood SES models). Model 1* is identical to Model 1 but replaces the biomarkers with measured aSES or cSES. For chronic conditions, a Poisson model was used; for the cardiometabolic conditions index (CMCI) and self-reported health status (SRHS) linear models were used; for mortality and dementia logistic models were used. 95% confidence intervals in parentheses. IRR, incidence rate ratio; OR, odds ratio. Sample sizes: number of chronic conditions=3120; CMCI=3120; SRHS=3118; mortality=3527; dementia=3120.

**Table S9.** Correlations between SES indices, SES biomarkers, and epigenetic aging measures in MESA

| | Adult SES index biomarker 450k (aSES-BIO450) | Childhood SES index biomarker 450k (cSES-BIO450) | Adult SES index (aSES) | Childhood SES index (cSES) | GrimAge | Dunedin-PACE |
|---|---|---|---|---|---|---|
| Adult SES index biomarker 450k (aSES-BIO450) | 1.000 | | | | | |
| Childhood SES index biomarker 450k (cSES-BIO450) | **0.122** | 1.000 | | | | |
| Adult SES index (aSES) | **0.205** | 0.008 | 1.000 | | | |
| Parental education | **0.194** | **0.068** | **0.403** | 1.000 | | |
| GrimAge | **0.356** | -0.010 | **0.079** | **0.120** | 1.000 | |
| DunedinPACE | **0.596** | 0.018 | **0.216** | **0.152** | **0.406** | 1.000 |

**Notes:** Pearson correlation coefficients with p<.05 under H0: r=0 are bolded. N=1,182.

**Table S10.** Associations between SES biomarkers and health outcomes in MESA

| | Adult SES | | | | | | | | |
|---|---|---|---|---|---|---|---|---|---|
| | | aSES | aSES-BIO450 | | | | | | |
| | Estimate Type | Model 1* | Model 1 | Model 2 | Model 3a | Model 4a | Model 3b | Model 4b | Model 5 |
| Cardiometabolic conditions index | Beta | 0.03 (-0.05, 0.11) | 0.14** (0.07, 0.22) | 0.14** (0.07, 0.22) | 0.10** (0.02, 0.19) | 0.10** (0.02, 0.19) | 0.04 (-0.04, 0.13) | 0.04 (-0.04, 0.13) | 0.14** (0.06, 0.21) |
| Self-reported health status | Beta | 0.26*** (0.19, 0.33) | 0.09** (0.03, 0.16) | 0.08** (0.02, 0.14) | 0.04 (-0.03, 0.12) | 0.05 (-0.02, 0.12) | 0.02 (-0.05, 0.10) | 0.02 (-0.05, 0.09) | 0.09** (0.02, 0.15) |
| Mortality | OR | 1.15 (0.94, 1.41) | 1.43** (1.19, 1.71) | 1.42** (1.18, 1.70) | 1.40** (1.14, 1.73) | 1.40** (1.14, 1.73) | 1.25* (1.00, 1.56) | 1.25* (1.00, 1.56) | 1.42** (1.18, 1.70) |
| ICD-based all cause dementia | OR | 1.11 (0.82, 1.51) | 0.85 (0.65, 1.12) | 0.85 (0.65, 1.11) | 0.80 (0.59, 1.09) | 0.80 (0.59, 1.09) | 0.80 (0.57, 1.11) | 0.80 (0.57, 1.11) | 0.87 (0.67, 1.14) |
| | Childhood SES | | | | | | | | |
| | | cSES | cSES-BIO450 | | | | | | |
| | Estimate Type | Model 1* | Model 1 | Model 2 | Model 3a | Model 4a | Model 3b | Model 4b | Model 5 |
| Cardiometabolic conditions index | Beta | 0.06* (-0.01, 0.14) | 0.02 (-0.05, 0.08) | 0.02 (-0.05, 0.08) | 0.02 (-0.05, 0.08) | 0.02 (-0.05, 0.08) | 0.02 (-0.04, 0.08) | 0.02 (-0.04, 0.08) | 0.02 (-0.05, 0.08) |
| Self-reported health status | Beta | 0.08** (0.02, 0.15) | 0.03 (-0.03, 0.09) | 0.02 (-0.03, 0.08) | 0.03 (-0.03, 0.09) | 0.02 (-0.03, 0.08) | 0.03 (-0.02, 0.09) | 0.03 (-0.03, 0.08) | 0.03 (-0.03, 0.08) |
| Mortality | OR | 1.12 (0.92, 1.37) | 0.92 (0.77, 1.10) | 0.90 (0.75, 1.08) | 0.91 (0.76, 1.09) | 0.89 (0.74, 1.08) | 0.93 (0.77, 1.11) | 0.91 (0.76, 1.09) | 0.92 (0.77, 1.10) |
| ICD-based all cause dementia | OR | 0.90 (0.68, 1.19) | 1.22* (0.96, 1.54) | 1.20 (0.95, 1.53) | 1.22 (0.96, 1.54) | 1.20 (0.95, 1.53) | 1.22 (0.96, 1.54) | 1.21 (0.95, 1.53) | 1.22* (0.96, 1.53) |

**Notes:** Table displays estimates from separate regressions of health or mortality on aSES-BIO450 or cSES-BIO450. The biomarkers were standardized for analysis. Model 1 controls for age, sex, race, current smoking, former smoking, and *APOE* e4 carrier status (dementia models only). Model 2 adds controls for measured aSES (in aSES-BIO450 regressions) or cSES (in cSES-BIO450 regressions). Models 3a and 4a add GrimAge to Model 1 (3a) or Model 2 (4a). Models 3b and 4b add DunedinPACE to Model 1 (3b) or Model 2 (4b). Model 5 adds measured cSES (for adult SES models) or measured aSES (for childhood SES models). Model 1* is identical to Model 1 but replaces the biomarkers with measured aSES or cSES. For cardiometabolic conditions index (CMCI) and self-reported health status (SRHS) linear models were used; for mortality and dementia logistic models were used. 95% confidence intervals in parentheses. OR, odds ratio. Sample sizes: CMCI=827; SRHS=848; Mortality=1180; Dementia=1180.

**Table S11.** Genomic feature enrichment for SES index biomarkers using two-sided Fisher's exact test

| | **aSES-BIO** | | **cSES-BIO** | | **aSES-BIO450** | |
|---|---|---|---|---|---|---|
| | Odds Ratio | P-value | Odds Ratio | P-value | Odds Ratio | P-value |
| DHS | **4.98** | **0.003** | 2.04 | 0.31 | 2.16 | 0.12 |
| Enhancer | **3.99** | **0.048** | 4.19 | 0.09 | 4.27 | 0.09 |
| Promoter | 1.28 | 0.64 | **3.28** | **0.018** | 1.20 | 0.69 |
| Shore/shelf | 1.38 | 0.49 | 1.76 | 0.26 | 1.23 | 0.56 |
| CpG Island | 1.95 | 0.13 | 2.49 | 0.10 | 1.28 | 0.55 |
| eQTM | **9.67** | **1.10E-05** | 8.56 | **8.30E-04** | **21.71** | **2.72E-14** |

**Notes:** DHS, DNase hypersensitive site; eQTM, expression quantitative trait methylation site. This table displays results from genomic feature enrichment analysis performed on the sets of CpGs selected for aSES-BIO and cSES- BIO by elastic net. An odds ratio > 1 indicates enrichment while an odds ratio <1 indicates depletion. Estimates with p<.05 are bolded.

**Table S12:** Gene Ontology Biological Processes Enrichment Analysis of aSES-BIO using eQTM from Keshawarz et al. (2023) (FDR q<0.01)

| ID | Description | BgRatio | Count | P-value | FDR q-value | Genes |
|---|---|---|---|---|---|---|
| GO:0008284 | positive regulation of cell population proliferation | 359/6537 | 30 | 7.93E-08 | 0.00019 | *NOG, LEF1, IL6ST, PPP1R16B, FLT3LG, LTBP3, MYC, LDLRAP1, EPHA1, PRKCA, NFATC2, JAK3, ADM, NCKAP1L, ST8SIA1, TNFRSF4, CD28, EPHA4, ITGB1, SLAMF1, GPR183, CCND2, RTKN2, CD248, WNT7A, FOXP3, IL2RA, HSF4, SEMA5A, IL7R* |
| GO:0030217 | T cell differentiation | 157/6537 | 17 | 2.04E-06 | 0.00017 | *CCR6, CCR7, IKZF3, RORC, LEF1, CAMK4, TCF7, BCL11B, RUNX3, JAK3, NCKAP1L, CD28, GPR183, GATA3, FOXP3, IL2RA, IL7R* |
| GO:0060070 | canonical Wnt signaling pathway | 124/6537 | 15 | 2.08E-06 | 0.0017 | *NOG, NKD1, LEF1, GPRC5B, EDA, DACT1, TCF7, MLLT3, IGFBP4, FUZ, KREMEN1, GATA3, WNT10A, WNT7A, SEMA5A* |
| GO:0035710 | CD4-positive, alpha-beta T cell activation | 70/6537 | 11 | 3.88E-06 | 0.0024 | *RORC, LEF1, NKG7, RUNX3, JAK3, NCKAP1L, CD28, GPR183, GATA3, FOXP3, IL2RA* |
| GO:0042475 | odontogenesis of dentin-containing tooth | 28/6537 | 7 | 9.97E-06 | 0.0040 | *EDAR, LEF1, EDA, BCL11B, ADM, FAM20A, WNT10A* |
| GO:0060548 | negative regulation of cell death | 405/6537 | 28 | 9.97E-06 | 0.0040 | *NOG, CCR7, LTK, BAG3, NPAS2, LEF1, IL6ST, RNF157, CHMP7, ITGA6, MYC, BCL11B, PRKCA, SOCS3, JAK3, CHMP1B, ANGPT1, NCKAP1L, CD28, BIRC3, ITGB1, GATA3, CCND2, RTKN2, WNT7A, SEMA5A, NEFL, IL7R* |
| GO:0198738 | cell-cell signaling by wnt | 170/6537 | 16 | 2.50E-05 | 0.0047 | *TRABD2A, NOG, NKD1, LEF1, GPRC5B, EDA, DACT1, TCF7, MLLT3, IGFBP4, FUZ, KREMEN1, GATA3, WNT10A, WNT7A, SEMA5A* |
| GO:0030334 | regulation of cell migration | 401/6537 | 26 | 6.47E-05 | 0.0087 | *CCR6, NOG, CCR7, ADTRP, ACE, NKD1, MMP28, LEF1, ITGA6, CXCR3, ZNF609 EPHA1, PRKCA, FUZ, PLEKHG3, ANGPT1, NCKAP1L, EPHA4, ITGB1, SLAMF1, TRADD, GPR183, GATA3, GCSAM, WNT7A, SEMA5A* |

**Notes:** aSES-BIO, adult socioeconomic status index biomarker; eQTM, expression quantitative trait methylation site; GO, Gene Ontology; BgRatio, Background Gene Ratio
This table displays results from GO Biological Processes gene-set analysis using eQTM data from Keshawarz et al. (2023) to test for enrichment of genes whose expression mapped to CpG sites included in the aSES-BIO. BgRatio represents the number of genes in the gene set that are part of the GO pathway divided by the total number of genes that have GO annotations in the gene set. Count represents the number of genes enriched to this GO entry from the input gene list. FDR q-value represents the P-value after FDR correction.

**Table S13:** Gene Ontology Biological Process Enrichment Analysis of cSES-BIO using eQTM from Keshawarz et al. (2023) (FDR q<0.01)

| ID | Description | BgRatio | Count | P-value | FDR q-value | Genes |
|---|---|---|---|---|---|---|
| GO:0006935 | chemotaxis | 258/6537 | 13 | 2.69E-07 | 0.00019 | *KLRK1, CCR5, CCL5, NRCAM, CCR4, EPHA4, CCR8, GPR183, SLAMF1, NOG, MMP28, GATA3, KIF5C* |
| GO:0042330 | taxis | 258/6537 | 13 | 2.69E-07 | 0.00019 | *KLRK1, CCR5, CCL5, NRCAM, CCR4, EPHA4, CCR8, GPR183, SLAMF1, NOG, MMP28, GATA3, KIF5C* |
| GO:0042110 | T cell activation | 306/6537 | 13 | 1.89E-06 | 0.00087 | *KLRK1, LAG3, EOMES, CD8B, CCL5, CD8A, CRTAM, GPR183, RORC, SLAMF1, TNFRSF4, CD28, GATA3* |
| GO:0071674 | mononuclear cell migration | 101/6537 | 8 | 2.49E-06 | 0.00087 | *KLRK1, CCR5, CCL5, CRTAM, GPR15, GPR183, SLAMF1, GATA3* |
| GO:0002708 | positive regulation of lymphocyte mediated immunity | 80/6537 | 7 | 5.74E-06 | 0.0015 | *KLRK1, LAG3, CRTAM, SLAMF1, CD226, CD28, GATA3* |
| GO:0002252 | immune effector process | 350/6537 | 13 | 8.35E-06 | 0.0018 | *KLRK1, LAG3, EOMES, CD8A, CRTAM, GPR183, RORC, SLAMF1, CD226, TNFRSF4, CD28, ACE, GATA3* |
| GO:0002407 | dendritic cell chemotaxis | 16/6537 | 4 | 9.38E-06 | 0.0018 | *CCR5, CCL5, GPR183, SLAMF1* |
| GO:0043583 | ear development | 63/6537 | 6 | 1.75E-05 | 0.0025 | *EPHA4, MAF, DDR1, NOG, GATA3, TTC39C* |
| GO:0045954 | positive regulation of natural killer cell mediated cytotoxicity | 19/6537 | 4 | 1.96E-05 | 0.0026 | *KLRK1, LAG3, CRTAM, CD226* |
| GO:0030098 | lymphocyte differentiation | 222/6537 | 10 | 2.08E-05 | 0.0026 | *LAG3, EOMES, CD8A, CRTAM, ITGB1, GPR183, RORC, SLAMF1, CD28, GATA3* |
| GO:0061564 | axon development | 181/6537 | 9 | 2.59E-05 | 0.0031 | *NRCAM, NEFL/EPHA4, ITGB1, TRIM46, DDR1, NOG, GATA3, KIF5C* |
| GO:0040012 | regulation of locomotion | 431/6537 | 13 | 7.59E-05 | 0.0064 | *KLRK1, CCL5, CCR4, EPHA4, TRADD, ITGB1, GPR183, PRAG1, SLAMF1, NOG, MMP28, ACE, GATA3* |
| GO:0010719 | negative regulation of epithelial to mesenchymal transition | 12/6537 | 3 | 1.38E-04 | 0.0097 | *EPHA4, NOG, GATA3* |

**Notes:** cSES-BIO, childhood socioeconomic status index biomarker; eQTM, expression quantitative trait methylation site; GO, Gene Ontology; BgRatio, Background Gene Ratio This table displays results from GO Biological Processes gene-set analysis using eQTM data from Keshawarz et al. (2023) to test for enrichment of genes whose expression mapped to CpG sites included in the cSES-BIO. BgRatio represents the number of genes in the gene set that are part of the GO pathway divided by the total number of genes that have GO annotations in the gene set. Count represents the number of genes enriched to this GO entry from the input gene list. FDR q-value represents the P-value after FDR correction.

**Table S14:** KEGG Pathway Gene Set Enrichment Analysis of cSES-BIO using eQTM from Keshawarz et al. (2023) (FDR q<0.01)

| ID | Description | BgRatio | Count | P-value | FDR q-value | Genes |
|---|---|---|---|---|---|---|
| hsa04514 | Cell adhesion molecules | 93/3232 | 7 | 9.41E-06 | 0.00096 | *5133, 926, 925, 4897, 3688, 10666, 940* |

**Notes:** cSES-BIO, childhood socioeconomic status index biomarker; eQTM, expression quantitative trait methylation site; KEGG, Kyoto Encyclopedia of Genes and Genomes; BgRatio, Background Gene Ratio. This table displays results from KEGG pathway analysis using eQTM data from Keshawarz et al. (2023) to test for enrichment of genes whose expression mapped to CpG sites included in the aSES-BIO. BgRatio represents the number of genes in the gene set that are part of the KEGG pathway divided by the total number of genes that have KEGG annotations in the gene set. Count represents the number of genes enriched to this KEGG entry from the input gene list. FDR q-value represents the P-value after FDR correction.

**Table S15:** Gene Ontology Biological Process Gene Set Enrichment Analysis of cSES-BIO450 using eQTM from Yao et al. (2021) (FDR q<0.01)

| ID | Description | BgRatio | Count | P-value | FDR q-value | Genes |
|---|---|---|---|---|---|---|
| GO:0002228 | natural killer cell mediated immunity | 34/2904 | 12 | 2.70E-09 | 5.45E-06 | *CD2, SH2D1B, KLRC4, KLRK1, GZMB, SLAMF7, KLRC4, KLRK1, NKG7, KLRD, KLRC3, KLRC1, KLRC2* |
| GO:0030101 | natural killer cell activation | 31/2904 | 11 | 1.22E-08 | 1.21E-05 | *CD2, KLRK1, SLAMF7, KLRC4, KLRK1, NKG7, KLRD1, PRDM1, TOX, KLRC3, KLRC1, KLRC2* |
| GO:0002250 | adaptive immune response | 184/2904 | 23 | 5.45E-07 | 2.75E-04 | *CD8B, CD8A, EOMES, TRGC2, SH2D1B, KLRK1, LEF1, TRGV2, TXK, SLAMF7, KLRC4, KLRK1, RORA, KLRD1, BACH2, PRDM1, CD84, MCOLN2, RFTN1, IL6ST, CD79A, CCR6, KLRC1, KLRC2* |
| GO:0002717 | positive regulation of natural killer cell mediated immunity | 16/2904 | 7 | 1.22E-06 | 4.10E-04 | *SH2D1B, KLRC4, KLRK1, KLRC4-KLRK1, KLRD1, KLRC3, KLRC2* |
| GO:0002252 | immune effector process | 206/2904 | 23 | 4.13E-06 | 0.0012 | *CD2, CD8A, EOMES, SH2D1B, KLRC4, KLRK1, GZMB, LEF1, A2M, SLAMF7, KRC4, KLRK1, RORA, NKG7, KLRD1, CFH, CFHR1, CD84, RFTN1, CD22, CCR6, KLRC3, KLRC1, KLRC2* |
| GO:0001909 | leukocyte mediated cytotoxicity | 58/2904 | 11 | 1.30E-05 | 0.0022 | *CD2, KLRC4, KLRK1, GZMB, SLAMF7, KLRC4, KLRK1, NKG7, KLRD1, KLRC3, KLRC1, KLRC2* |
| GO:0001906 | cell killing | 69/2904 | 12 | 1.31E-05 | 0.0022 | *CD2, KLRC4, KLRK1, GZMB, SLAMF7, KLRC4, KLRK1, NKG7, KLRD1, CFH, KLRC3, KLRC1, KLRC2* |
| GO:0001775 | cell activation | 326/2904 | 29 | 1.95E-05 | 0.0030 | *CD8B, CD2, CD8A, EOMES, TRGC2, SH2D1B, KLRK1, CCR7, LEF1, CCL5, TXK, SLAMF7, KLRC4, KLRK1, RORA, NKG7, KLRD1, ITGB1, ATM, PRDM1, CD84, IL6ST, TOX, CD22, CD79A, FCRL1, CCR6, KLRC3, KLRC1, KLRC2* |
| GO:0032814 | regulation of natural killer cell activation | 16/2904 | 6 | 2.17E-05 | 0.0031 | *KLRD1, PRDM1, TOX, KLRC3, KLRC1, KLRC2* |
| GO:0007399 | nervous system development | 383/2904 | 32 | 2.39E-05 | 0.0032 | *EOMES, MAL, MYO6, SERINC5, PTPRM, LEF1, RORA, GPRIN3, ITGB1, ADAM23, PTPN13, APBB1, SMARCA1, EPHA4, SLC4A7, LRP6, ATM, PRDM1, ITGA6, AUTS2, NOG, DACT1, APBA2, IGF1R, IL6ST, CCR4, TOX, NRCAM, KIF5C, CCDC141, LRRN3, P2RX5* |
| GO:0002449 | lymphocyte mediated immunity | 117/2904 | 15 | 4.81E-05 | 0.0051 | *CD2, CD8A, SH2D1B, KLRC4, KLRK1, GZMB, SLAMF7, KLRC4, KLRK1, NKG7, KLRD1, RFTN1, CCR6, KLRC3, KLRC1, KLRC2* |

**Notes:** cSES-BIO450, 450K derived childhood socioeconomic status index biomarker; eQTM, expression quantitative trait methylation site; GO, Gene Ontology; BgRatio, Background Gene Ratio. This table displays results from GO Biological Processes gene-set analysis using eQTM data from Yao et al. (2021) to test for enrichment of genes whose expression mapped to CpG sites included in the cSES-BIO450. BgRatio represents the number of genes in the gene set that are part of the GO pathway divided by the total number of genes that have GO annotations in the gene set. Count represents the number of genes enriched to this GO entry from the input gene list. FDR q-value represents the P-value after FDR correction.

**Figure S3:** Dotplot showing the Gene Ontology Biological Processes (BP) enrichment results for genes whose expression was associated with CpGs selected for aSES-BIO using eQTM from Keshawarz et al. (2023)

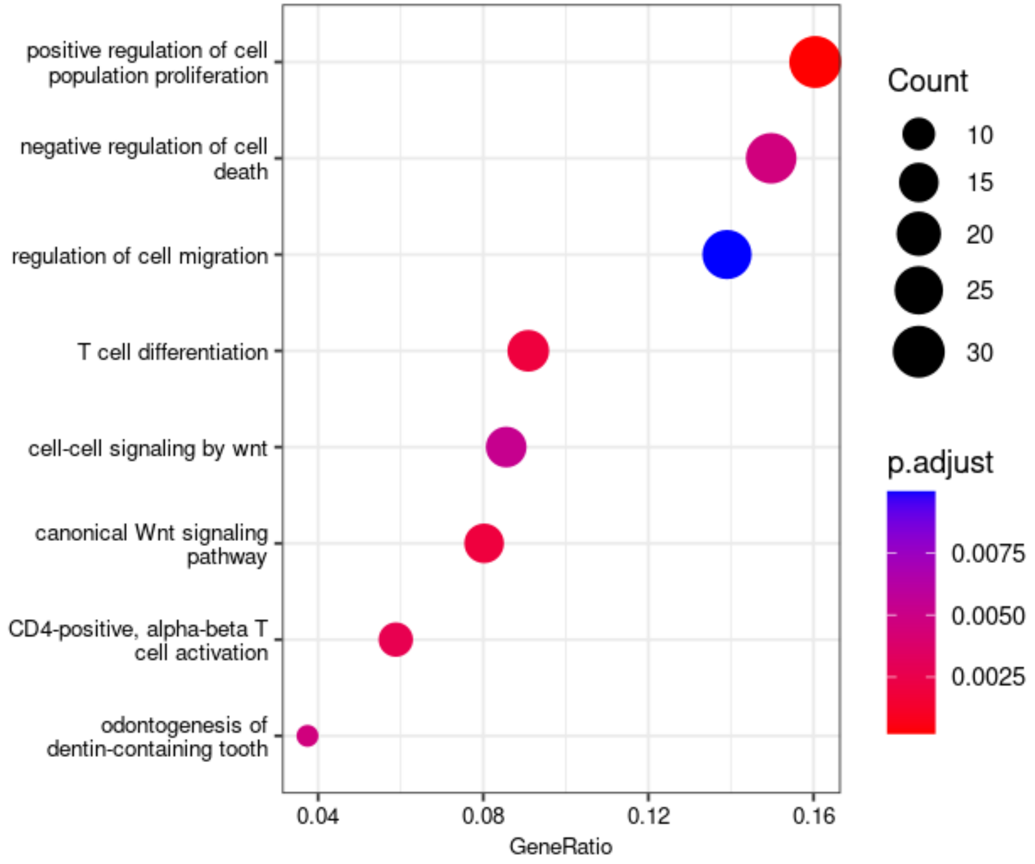


**Notes:** Dot size represents the number of genes in a specific GO category. Dot color represents the FDR q value, with a threshold of FDR q<0.01.

**Figure S4:** Dotplot showing the Gene Ontology Biological Processes (BP) genes whose expression was associated with CpGs selected for cSES-BIO using eQTM from Keshawarz et al. (2023)

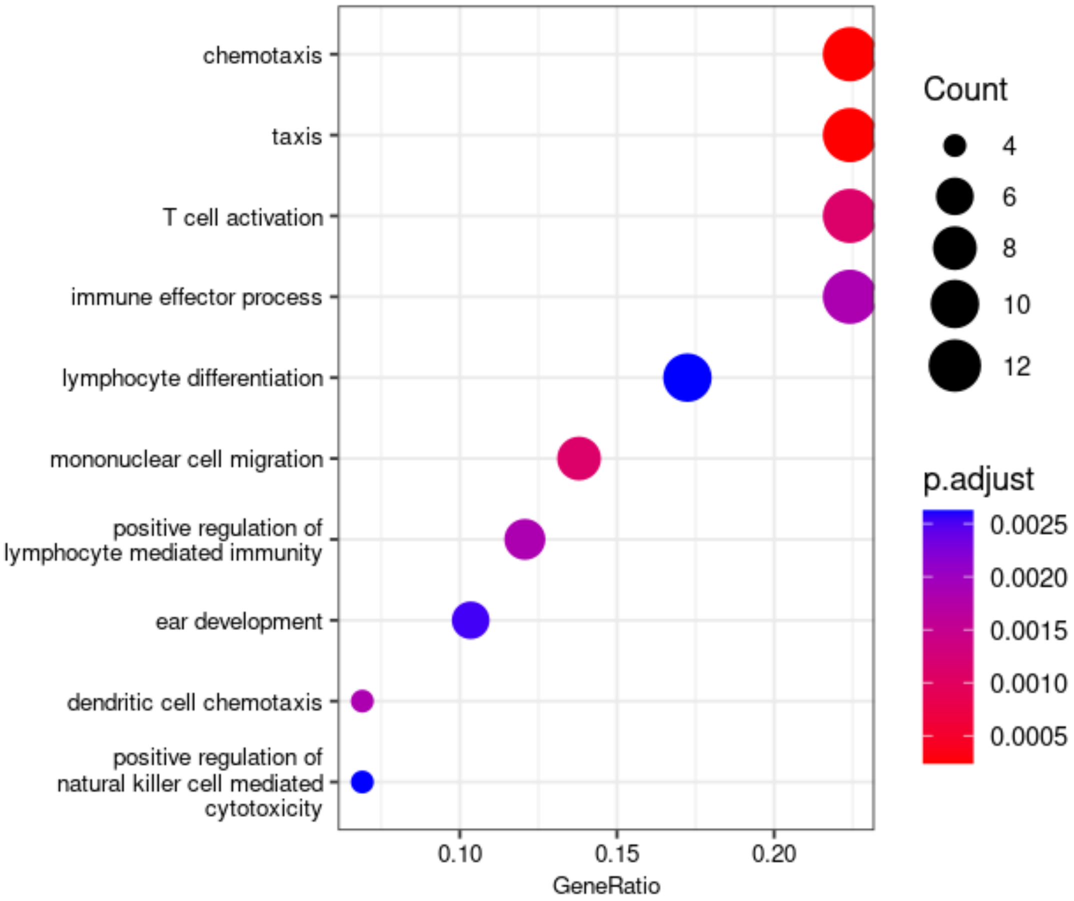


**Notes:** Dot size represents the number of genes in a specific GO category. Dot color represents the FDR q value, with a threshold of FDR q<0.01.

**Figure S5:** Dotplot showing the Gene Ontology Biological Processes (BP) enrichment results for genes whose expression was associated with CpGs selected for cSES-BIO450k using eQTM from Yao et al. (2021)

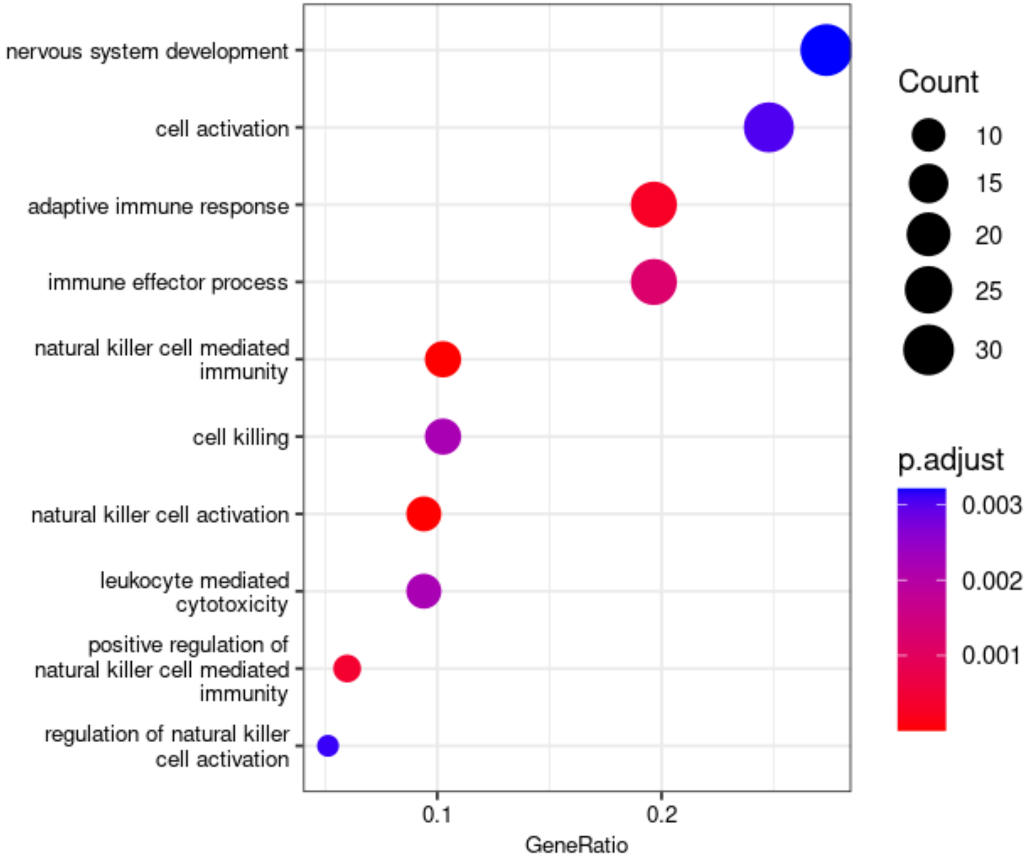


**Notes:** Dot size represents the number of genes in a specific GO category. Dot color represents the FDR q value, with a threshold of FDR q<0.01.

**Table S16.** Traits previously associated with CpG sites in the adult SES index biomarker (aSES-BIO)

| CpG | Chr. | Pos. | UCSC RefGene Name | Traits previously associated with CpG sites in EWAS |
|---|---|---|---|---|
| cg18181703 | 17 | 76354621 | *SOCS3* | Age, C-reactive protein, COPD, BMI, Smoking, Soluble tumor necrosis factor receptor 2, Crohn's disease, FEV1, Type 2 diabetes, Healthy eating, Inflammatory bowel disease, Chronic pain, Alcohol consumption, Cognitive abilities, ischemic heart disease, Primary Sjogrens syndrome, educational attainment, Stroke, Liver cirrhosis, Rheumatoid arthritis |
| cg11047325 | 17 | 76354934 | *SOCS3* | C-reactive protein, COPD, Ischemic heart disease, Smoking, Rheumatoid arthritis, Chronic pain, Type 2 diabetes, FEV1, Stroke, Liver cirrhosis, chronic kidney disease |
| cg21327712 | 1 | 111744503 | *DENND2D* | C-reactive protein, Type 2 diabetes, Chronic pain, Liver cirrhosis, COPD |
| cg01894508 | 2 | 70189111 | *ASPRV1* | C-reactive protein, COPD, Chronic pain, Diet quality, Schizophrenia, Ischemic heart disease, Type 2 diabetes, Age |
| cg18062721 | 3 | 11643427 | *VGLL4* | C-reactive protein, BMI, Alcohol consumption, Age, Type 2 diabetes, Crohn's disease |
| cg03362418 | 22 | 50965563 | *TYMP; SCO2* | Eosinophilia, COPD, C-reactive protein, Type 2 diabetes |
| cg25008217 | 19 | 41882654 | *TMEM91* | Atopy, COPD |
| cg17461390 | 19 | 17428411 | *DDA1* | C-reactive protein, Type 2 diabetes |
| cg02767093 | 13 | 99130655 | *STK24* | Smoking, Age, COPD, C-reactive protein, Type 2 diabetes |
| cg25145728 | 16 | 89628775 | *RPL13* | Eosinophilia, COPD, C-reactive protein |
| cg02017926 | 12 | 123754328 | *CDK2AP1* | Age, Sex, Clear cell renal carcinoma, C-reactive protein, COPD |
| cg10922280 | 16 | 68034227 | *DPEP2* | Age, C-reactive protein, BMI, Type 2 diabetes, Chron's disease, Cognitive abilities, Eosinophilia |
| cg10206344 | 16 | 31483277 | *TGFB1I1* | Age, C-reactive protein |
| cg11454468 | 3 | 142569221 | *PCOLCE2* | Eosinophilia, C-reactive protein, Type 2 diabetes, COPD |
| cg11183072 | 17 | 37894397 | *GRB7* | Age, Clear cell renal carcinoma, C-reactive protein, COPD |
| cg10842530 | 14 | 69438358 | *ACTN1* | C-reactive protein |
| cg03031609 | 10 | 7453871 | *SFMBT2* | Age |
| cg24445316 | 1 | 23889092 | | Age, Waist circumference, Sex |
| cg06864083 | 17 | 40832319 | *CCR10* | Age, Sex, C-reactive protein |
| cg03090734 | 10 | 44525757 | | C-reactive protein, COPD |
| cg10946295 | 2 | 119913307 | | Age, Sex |
| cg27637521 | 17 | 76355202 | *SOCS3* | Smoking, Age, BMI, C-reactive protein, Clear cell renal carcinoma, COPD, Type 2 diabetes |
| cg06021088 | 2 | 127822551 | *BIN1* | Age, Primary Sjogrens syndrome, Clear cell renal carcinoma, Pancreatic ductal adenocarcinoma, COPD, C-reactive protein |
| cg19574915 | 15 | 89195555 | *ISG20* | Age, C-reactive protein, Type 2 diabetes |
| cg08852765 | 6 | 33396407 | *SYNGAP1* | Age |

**Notes:** Chr., chromosome; EWAS, epigenome-wide association study; Pos., position; UCSC, University of California Santa Cruz Genome Browser. This table displays traits that have been previously associated with CpG sites selected for the aSES-biomarker in prior EWAS. All associations were identified in the MRC-IEU catalog of epigenome-wide association studies with p<1E-06.

**Table S17.** Traits previously associated with CpG sites in the childhood SES index biomarker (cSES-BIO)

| CpG | Chr. | Pos. | UCSC RefGene Name | Traits previously associated with CpG sites in EWAS |
|---|---|---|---|---|
| cg04887278 | 6 | 83903927 | *PGM3; RWDD2A* | Age, Atopy |
| cg03519157 | 11 | 46367033 | *DGKZ* | Age, Atopy |
| cg08469255 | 6 | 30851069 | *DDR1* | Age, Primary Sjorgren's syndrome, C-reactive protein |
| cg17810176 | 19 | 36035831 | *GAPDHS; TMEM147* | Age, Clear cell renal carcinoma |
| cg22505924 | 2 | 64488157 | | C-reactive protein, COPD, Chronic pain |
| cg27496526 | 15 | 41805530 | *LTK* | Age, Rheumatoid arthritis |
| cg25563256 | 17 | 7341641 | *FGF11* | Age, Sex, COPD |
| cg16329896 | 7 | 47515060 | *TNS3* | Atopy, Age, Primary Sjorgren's syndrome |
| cg10394832 | 1 | 111889533 | *C1orf88* | Age, C-reactive protein |
| cg17496659 | 1 | 3568245 | *TP73* | Age, Clear cell renal carcinoma, Atopy, HIV infection, COPD, BMI |
| cg21159993 | 5 | 139554405 | *C5orf32* | Age, Clear cell renal carcinoma, Atopy |
| cg19989295 | 14 | 24641077 | *REC8* | Age, Clear cell renal carcinoma |
| cg13825574 | 9 | 140117257 | *RNF208* | Age |
| cg05302489 | 6 | 31760426 | *VARS* | Smoking, Educational attainment, Age, COPD, C-reactive protein, Chronic pain, ischemic heart disease |
| cg02170695 | 21 | 38909809 | | --- |
| cg13067434 | 15 | 63836865 | *USP3* | C-reactive protein |
| cg04382191 | 3 | 9437901 | *SETD5; THUMPD3-AS1* | -- |

**Notes:** Chr., chromosome; EWAS, epigenome-wide association study; Pos., position; UCSC, University of California Santa Cruz Genome Browser. This table displays traits that have been previously associated with CpG sites selected for the cSES-biomarker. All associations were identified in the MRC-IEU catalog of epigenome-wide association studies with p<1E-06.

**Table S18.** Traits previously associated with CpG sites in the 450K derived adult SES index biomarker (aSES-BIO450)

| CpG | Chr. | Pos. | UCSC RefGene Name | Traits previously associated with CpG sites in EWAS |
|---|---|---|---|---|
| cg18181703 | 17 | 76354621 | *SOCS3* | Age, C-reactive protein, COPD, BMI, Smoking, Soluble tumor necrosis factor receptor 2, Crohn's disease, FEV1, Type 2 diabetes, Healthy eating, Inflammatory bowel disease, Chronic pain, Alcohol consumption, Cognitive abilities, ischemic heart disease, Primary Sjogrens syndrome, educational attainment, Stroke, Liver cirrhosis, Rheumatoid arthritis |
| cg01894508 | 2 | 70189111 | *ASPRV1* | C-reactive protein, COPD, Chronic pain, Diet quality, Schizophrenia, Ischemic heart disease, Type 2 diabetes, Age |
| cg18062721 | 3 | 11643427 | *VGLL4* | C-reactive protein, BMI, Alcohol consumption, Age, Type 2 diabetes, Crohn's disease |
| cg01406381 | 19 | 47288263 | *SLC1A5* | Clear cell renal carcinoma, COPD, C-reactive protein, Type 2 diabetes, Smoking, Eosinophilia, Ischemic heart disease, Primary Sjogrens syndrome, Cognitive abilities, Sex |
| cg25691553 | 3 | 49057884 | *NDUFAF3; DALRD3; MIR425* | Age, COPD, C-reactive protein, Smoking, Stroke, Type 2 diabetes, ischemic heart disease |
| cg02767093 | 13 | 99130655 | *STK24* | Smoking, Age, COPD, C-reactive protein, Type 2 diabetes |
| cg24704287 | 19 | 13951481 | | Clear cell renal carcinoma, COPD, C-reactive protein, Smoking, Type 2 diabetes, Primary Sjogrens syndrome, Soluble tumor necrosis factor receptor 2, Educational attainment, chronic kidney disease |
| cg19769182 | 16 | 29823868 | *PRRT2* | Age, C-reactive protein |
| cg02017926 | 12 | 123754328 | *CDK2AP1* | Age, Sex, Clear cell renal carcinoma, C-reactive protein, COPD |
| cg08696931 | 12 | 123754071 | *CDK2AP1* | Age, Sex, BMI, C-reactive protein, COPD |
| cg08469255 | 6 | 30851069 | *DDR1* | Age, C-reactive protein, Primary Sjogrens syndrome, |
| cg10922280 | 16 | 68034227 | *DPEP2* | Age, C-reactive protein, BMI, Type 2 diabetes, Chron's disease, Cognitive abilities, Eosinophilia |
| cg10206344 | 16 | 31483277 | *TGFB1I1* | Age, C-reactive protein |
| cg25130381 | 1 | 27440721 | *SLC9A1* | C-reactive protein, Age, HDL cholesterol, BMI, COPD, Primary Sjogrens syndrome, Type 2 diabetes, Chron's disease |
| cg11183072 | 17 | 37894397 | *GRB7* | Age, Clear cell renal carcinoma, C-reactive protein, COPD |
| cg01101459 | 1 | 234871477 | | Age, C-reactive protein, Type 2 diabetes, BMI, HDL cholesterol, COPD, Chronic pain, Diet quality, Soluble tumor necrosis factor receptor 2, Chron's disease |
| cg08136809 | 19 | 41882642 | *TMEM91* | Age, Sex, Clear cell renal carcinoma, Smoking, COPD, C-reactive protein |
| cg11846112 | 1 | 227729906 | | Age, C-reactive protein, Atopy, COPD |
| cg03031609 | 10 | 7453871 | *SFMBT2* | Age |
| cg18385254 | 18 | 77724395 | *HSBP1L1* | Age |
| cg27491190 | 12 | 46767943 | *SLC38A2* | Age, Alcohol consumption |
| cg06864083 | 17 | 40832319 | *CCR10* | Age, Sex, C-reactive protein |
| cg26504305 | 19 | 19281019 | *LOC729991-MEF2B; MEF2B* | Age, Sex |
| cg27637521 | 17 | 76355202 | *SOCS3* | Smoking, Age, BMI, C-reactive protein, Clear cell renal carcinoma, COPD, Type 2 diabetes |

| | | | | |
|---|---|---|---|---|
| cg22488164 | 12 | 14716910 | *PLBD1* | C-reactive protein, BMI, Age, HDL cholesterol, Fasting insulin, Sex, Alcohol consumption, Chronic pain, COPD |
| cg15866367 | 13 | 49796387 | *MLNR* | Age, Sex, C-reactive protein, Chronic pain, COPD, Smoking |
| cg06021088 | 2 | 127822551 | *BIN1* | Age, Primary Sjogrens syndrome, Clear cell renal carcinoma, Pancreatic ductal adenocarcinoma, COPD, C-reactive protein |
| cg19574915 | 15 | 89195555 | *ISG20* | Age, C-reactive protein, Type 2 diabetes |
| cg08852765 | 6 | 33396407 | *SYNGAP1* | Age |

**Notes:** Chr., chromosome; EWAS, epigenome-wide association study; Pos., position; UCSC, University of California Santa Cruz Genome Browser. This table displays traits that have been previously associated with CpG sites selected for the 450K derived aSES-biomarker. All associations were identified in the MRC-IEU catalog of epigenome-wide association studies with p<1E-06.

**Table S19.** Traits previously associated with CpG sites in the 450K derived childhood SES index biomarker (cSES-BIO450)

| CpG | Chr. | Pos. | UCSC RefGene Name | Traits previously associated with CpG sites in EWAS |
|---|---|---|---|---|
| cg04887278 | 6 | 83903927 | *PGM3; RWDD2A* | Age, Atopy |
| cg03519157 | 11 | 46367033 | *DGKZ* | Age, Atopy |
| cg08469255 | 6 | 30851069 | *DDR1* | Age, Primary Sjorgren's syndrome, C-reactive protein |
| cg17810176 | 19 | 36035831 | *GAPDHS; TMEM147* | Age, Clear cell renal carcinoma |
| cg27496526 | 15 | 41805530 | *LTK* | Age, Rheumatoid arthritis |
| cg25563256 | 17 | 7341641 | *FGF11* | Age, Sex, COPD |
| cg16329896 | 7 | 47515060 | *TNS3* | Atopy, Age, Primary Sjorgren's syndrome |
| cg12452298 | 15 | 67134587 | | Age, C-reactive protein |
| cg10394832 | 1 | 111889533 | *C1orf88* | Age, C-reactive protein |
| cg17496659 | 1 | 3568245 | *TP73* | Age, Clear cell renal carcinoma, Atopy, HIV infection, COPD, BMI |
| cg21159993 | 5 | 139554405 | *C5orf32* | Age, Clear cell renal carcinoma, Atopy |
| cg19989295 | 14 | 24641077 | *REC8* | Age, Clear cell renal carcinoma |
| cg13825574 | 9 | 140117257 | *RNF208* | Age |
| cg05302489 | 6 | 31760426 | *VARS* | Smoking, Educational attainment, Age, COPD, C-reactive protein, Chronic pain, ischemic heart disease |
| cg12748367 | 2 | 216301062 | *FN1* | --- |

**Notes:** Chr., chromosome; EWAS, epigenome-wide association study; Pos., position; UCSC, University of California Santa Cruz Genome Browser. This table displays traits that have been previously associated with CpG sites selected for the 450k derived cSES-biomarker. All associations were identified in the MRC-IEU catalog of epigenome-wide association studies with p<1E-06.
